# Disparities in ABO Blood Type Determination Across Diverse Ancestries: A Systematic Review and Validation in the *All of Us* Research Program

**DOI:** 10.1101/2024.02.21.24302372

**Authors:** Kiana L. Martinez, Andrew Klein, Jennifer R. Martin, Chinwuwanuju U. Sampson, Jason B. Giles, Madison L. Beck, Krupa Bhakta, Gino Quatraro, Juvie Farol, Jason H. Karnes

**Author notes:** Correspondence to: Jason H. Karnes, PharmD, PhD Associate Professor, University of Arizona R. Ken Coit College of Pharmacy 1295 N Martin AVE, Tucson, AZ 85721.

## Abstract

**Background:** ABO blood types have widespread clinical use and robust associations with cardiovascular disease. Many studies determine ABO blood types using tag single nucleotide polymorphisms (tSNPs) to characterize functional variation. However, tSNPs with low linkage disequilibrium (LD) may promote misinference of ABO blood types, particularly in diverse populations.

**Methods:** Bibliographic databases were searched for studies (2005-2022) using tSNPs to determine ABO alleles in accordance with PRISMA 2020 guidelines. We calculated linkage between tSNPs and functional variants across inferred continental ancestry groups from 1000 Genomes (AFR, AMR, EAS, EUR). We compared r^2^ across ancestry and assessed real-world consequences by comparing tSNP-derived blood types to serology in a large, diverse population from the *All of Us* Research Program (AoURP).

**Results:** We observe a lack of phasing and frequent use of inappropriate tSNPs in blood type determination, particularly for O alleles. Linkage between functional variants and O allele tSNPs was significantly lower in African (median r^2^=0.443) compared to East Asian (r^2^=0.946, p=1.1×10-5) and European (r^2^=0.869, p=0.023). In AoURP, discordance between tSNP-derived blood types and serology was high across all SNPs in African ancestry individuals and linkage was strongly correlated with discordance across all ancestries (ρ=-0.90, p=3.08×10-23).

**Conclusion:** We observe common use of inappropriate tSNPs to determine ABO blood type, particularly for O alleles and with some tSNPs mistyping up to 58% of individuals. Our results highlight the lack of transferability of tSNPs across ancestries and potential exacerbation of disparities in genomic research for underrepresented populations.

## Introduction

Patients with European ancestry make up 12% of the world’s population, but account for approximately 81% of individuals in genomic studies^1,2^. This lack of representation in genomic studies has led to multiple racial and ethnic disparities in genomic research. Among such disparities is the assumption that tag single-nucleotide polymorphisms (tSNPs) are appropriate proxies for functional variation across ancestry groups. Because of differences in minor allele frequency (MAF) and linkage disequilibrium (LD) structure, tSNPs are not always portable across ancestry groups and inappropriate use of tSNPs may lead to reduced statistical power and erroneous associations^3,4^. This practice may exacerbate existing disparities in genomic research for underrepresented populations, especially when important clinical traits are inferred based on tSNP variation.

ABO blood types have widespread clinical use, are among the first biomarkers tested in the newborn, and are robustly associated with myriad phenotypes, such as hemolytic disease of the newborn transfusion and solid organ transplant-related complications, and thromboembolic disease^5–9^. Individuals with non-O blood types have been shown to have increased risk for coronary artery disease, myocardial infarction, deep vein thrombosis, and stroke^10–13^. The four ABO blood groups (A, B, AB, and O) are largely the result of a small number of functional variants that produce combinations of ABO alleles (A, B, and O)^14–16^. While serological typing is considered the gold standard for determining ABO blood types, studies often determine ABO blood type using genetic data when serological typing results are not collected or not available^17,18^. These studies often use tSNPs to determine ABO blood type because important functional variation in ABO includes single base pair insertion/deletion polymorphisms that are not easily or accurately characterized on arrays or with genome-wide imputation^19^. Additionally, most ABO tSNPs used to identify blood groups were originally identified in European and East Asian populations^20,21^, leading to the possibility that tSNPs are used inappropriately in diverse populations^4^.

Many diverse populations, including those with admixed American and African ancestry, are included in studies that infer ABO blood groups based on genetic variants^22–25^. However, many of the commonly utilized tSNPs in ABO are unlikely to accurately capture functional variation in these populations due to differences in LD patterns and MAFs ^21,26^. In this study, our primary aim is to evaluate the portability and suitability of tSNPs used to determine ABO alleles and blood types across diverse populations in published literature. We aimed to evaluate inconsistencies in racial/ethnic representation and bioinformatic methodologies across published studies investigating genotype-derived ABO blood groups. Finally, we investigated the real-world consequences of utilization of tSNPs from existing studies through examination of LD patterns, genotype-derived ABO blood groups, and ABO serological test results from the diverse participants in the *All of Us* Research Program (AoURP).

## Methodology

In the systematic review portion of this study, we captured all recent publications (publication dates 2005-2022) that used single-nucleotide polymorphisms (SNPs) to determine ABO alleles (A, B, and O). ABO alleles are derived from pre-defined functional variants and are used to determine ABO blood types: OO (O blood type); AA, AO (A blood type); BB, BO (B blood type); AB (AB blood type). We identified and recorded SNPs, either functional variants or tSNPs, that were used to determine ABO alleles. As studies did not report LD between their utilized tSNPs and functional variants within their population, we calculated LD using r^2^ in reference populations provided by the 1000 Genomes Project (1KGP)^27^. Because studies also rarely reported genetically-derived ancestry for their populations, we inferred continental ancestry from 1KGP superpopulations (African ancestry [AFR], Admixed American ancestry [AMR], East Asian ancestry [EAS], European ancestry [EUR], and South Asian ancestry [SAS]) to estimate r^2^ between functional variants and tSNPs. To evaluate the real-life consequences of utilizing potentially inappropriate tSNPs in place of functional variants, we compared ABO blood types and alleles derived from tSNPs utilized in existing studies, functional variants, and ABO serology test results in the diverse population with whole genome sequencing from the AoURP^28^. We calculated discordance between these different derivations of ABO blood types and alleles across ancestries in the AoURP. Approval for human subjects research was not required since all data used in this study was publicly available. Research conducted within the AoURP is considered non-human subjects research since all data were de-identified and available for public access.

### Systematic Review Search Strategy, Inclusion Criteria, Data Extraction, and Derived Data

We conducted a systematic search for peer-reviewed publications (2005-2022) that used SNPs to determine ABO blood types **(Figure 1)**. The results of this review is reported in accordance with the Preferred Reporting Items for Systematic reviews and Meta-Analyses (PRISMA) 2020 statement and checklist^29^. Detailed descriptions of the search strategy, inclusion criteria, data extraction, and derived data are presented in the Supplemental Materials (See **Supplemental Methods** and **Tables S1-S6**). We utilized the reference management tool EndNote 20 (Clarivate, Philadelphia, PA, USA) for the initial compilation of references based on our search strategy, and then used DistillerSR (DistillerSR, Ottawa, Canada) as our literature reviewer software that enabled us to efficiently screen references and extract data. The following bibliographic databases were searched: PubMed/Medline (National Library of Medicine), Embase (Elsevier), Scopus (Elsevier), Ovid/Medline (Wolters Kluwer), and CINHAL Plus with Full Text (Ebsco). In addition, Dissertations and Theses Global (ProQuest) were also searched as well as Google Scholar. These searches were conducted within 1 week in August 2022. Key words including MeSH (Medical Subject Heading) terms and free-text words were searched in titles, abstracts, and text. The full complete queries conducted in each database are provided in the **Supplementary Materials** (**Table S2**).

**Figure 1.**
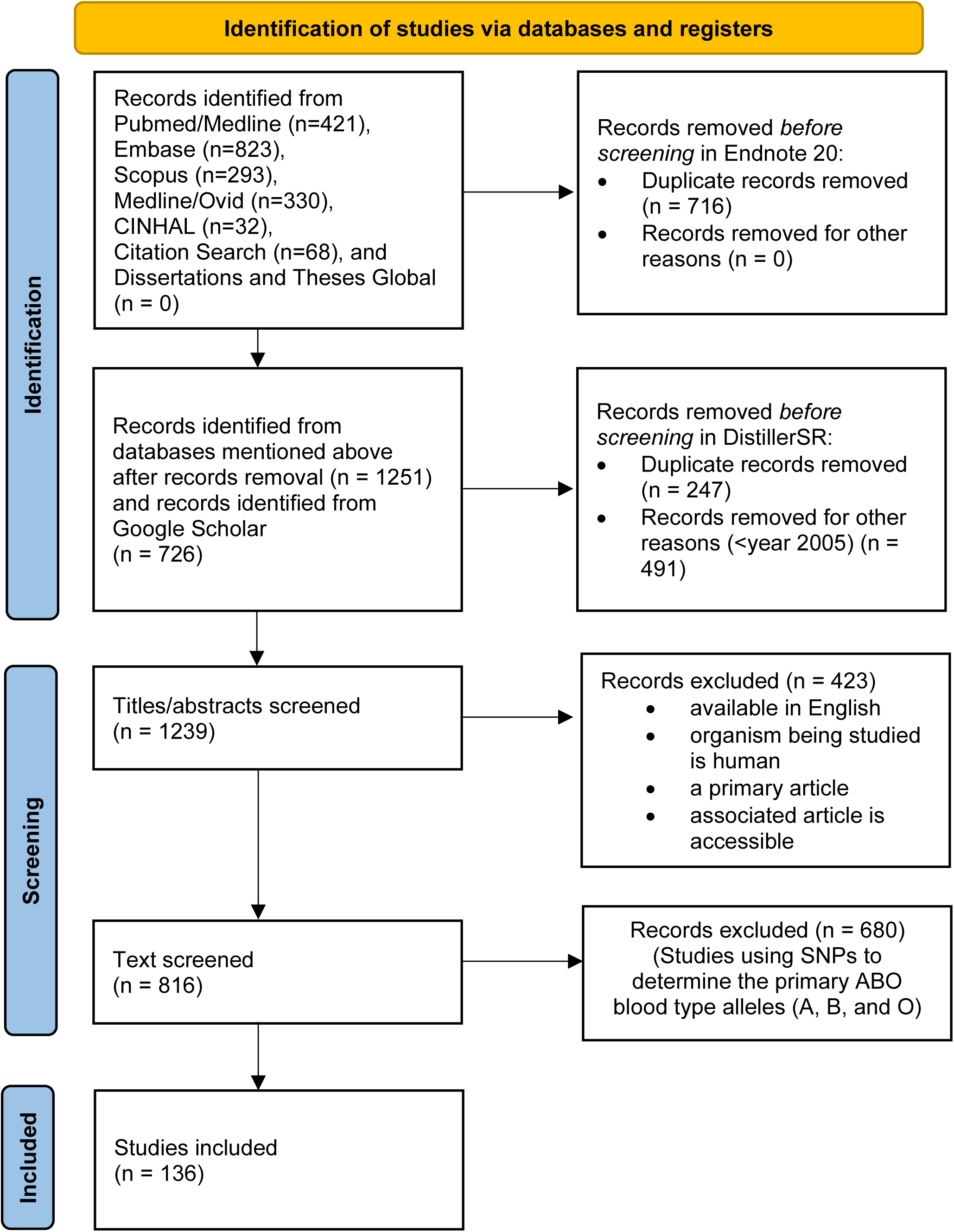
Study selection process per PRISMA 2020 guidelines.

The publications meeting the following criteria were included for the initial title and abstract evaluation: 1) available in the English language, 2) the organism being studied is human, and 3) is a primary article. In a secondary evaluation we screened the full text and included only peer-reviewed articles that used single SNPs to derive ABO alleles from genomic data. Major exclusion criteria were: 1) clinical trials, 2) reviews, and 3) meta-analysis. The screening forms are available in **Table S3** and **Table S4**. The bibliographic information associated with each reference, such as reference title, authors, and journal, were automatically extracted by DistillerSR. The primary data extracted included: 1) SNPs used for each ABO allele, 2) presence of a haplotype analysis, 3) cohort size, 4) cohort population description, 5) primary phenotype of interest, and 6) sequencing/genotyping platform (**Table S4**). We ensured consistency of the collected data by having our data independently extracted by two different reviewers with disagreement between reviewers being resolved by consensus.

To better understand the range of phenotypes assessed in conjunction with ABO blood type, we categorized the primary phenotype of the studies into twelve categories (hematological, cardiovascular, oncology, infections disease, digestive system/gastrointestinal, inflammatory, immune system, hepatic, neurological, metabolic, miscellaneous, multiple phenotypes). Because genotyping technology influences the functional ABO variation that could be determined, we also collected sequencing and genotyping platforms used to determine the SNPs and ABO alleles. We categorized these platforms into four groups: 1) sequencing, 2) genotyping, 3) PCR/specific target, and 4) multiple genotyping/sequencing platforms. Definitions for these categories are available in the **Supplemental Materials**. Since the use of phased genetic data and imputation of haplotype structure is required to accurately determine ABO alleles (**Table S5**), we also identified how many studies used a haplotype analysis to assign functional variants or tSNPs to individual copies of chromosome nine.

### Inferred Continental Ancestry for Assessment of LD

As our aim was to evaluate tSNPs across diverse populations, we were required to determine LD between tSNPs and functional variants, as well as describe reported populations in terms of continental ancestry. Since studies using tSNPs did not report population-specific LD values for linkage between functional variants and tSNPs, we estimated LD using r^2^ from representative populations from 1KGP^27^. We used LDLink^30^, specifically the LDpop^31^ module, to calculate r^2^ specific to super- and sub-populations as defined by 1KGP^27^. For our primary analysis, r^2^ was calculated for each of the 1KGP superpopulations (AFR, AMR, EAS, EUR, or SAS) that corresponded to each study population as described below. In studies where ABO functional variation was used, r^2^ was determined to be 1. In studies where more than one SNP was used and the population for which the SNP was used was not described, we assumed studies used the SNP with the highest r^2^ value for their population.

For each study in the systematic review, we inferred continental ancestry as described by the International Genome Sample Resource (IGSR) for their 1KGP^27^ phase three collection. The vast majority of studies included in this systematic review did not describe genetically-derived ancestry for their participants. For example, many studies described their populations using racial terminology such as “White” or “Caucasian” or in terms of the population’s geography such as being “Japanese” or “Finnish”. Thus, in scenarios where continental ancestry was not provided, we inferred continental ancestry based on these other descriptors and categorized them into AFR, AMR, EAS, EUR, or SAS.

### Statistical Analysis for Systematic Review

We visually examined LD structure of the reported SNPs across populations using LDBlockShow^32^ and data from 1KGP on GRCh38^27^. R^2^ was calculated between functional variants and all reported tSNPs for each superpopulation. A tSNP was only included in our population-specific analysis if that tSNP was actually used to derive ABO blood types in an existing study including a population with similar ancestry. To examine within and across population variation of LD, we calculated r^2^ between all reported tSNPs and functional variants across the 26 subpopulations described in 1KGP. Values were then grouped by superpopulation to evaluate variation within and across superpopulations. The functional variant for O vs non-O alleles was defined as rs8176719 and the functional variants for A vs B alleles were defined as rs7853989, rs8176743, rs8176746, and rs8176747. For our primary analyses, we calculated a single r^2^ value for each population in each study between the study’s reported tSNP and the functional variant (rs8176719 for O vs non-O and rs8176746 for A vs B) using the matching 1KGP superpopulation. We first conducted a Kruskal Wallis test for differences in median r^2^ across superpopulations for each tSNP using R^33^. Then we performed pairwise comparisons across superpopulations for each tSNP with Dunn’s test of multiple comparisons using rank sums. Our primary analysis excluded studies that used ABO allele-determining functional variants, however a secondary analysis was also performed that included studies using functional variants. We excluded a single study with inferred South Asian (SAS) continental ancestry due to the small sample size, restricting our inferred continental ancestries of interest to AFR, AMR, EAS and EUR. Studies that had populations with unclear ancestry were also excluded. We calculated the coefficient of variation for r^2^ to evaluate variance across superpopulations in a kernel density plot.

### Derivation of Population and Data for All of Us Research Program Analysis

To evaluate real-world consequences of utilizing tSNPs in place of functional variants or serology, we utilized a subset of AoURP^28^. The AoURP, sponsored by the National Institutes of Health (NIH), is a longitudinal cohort study with an aim to advance precision medicine by collecting electronic health records (EHR), participation-provided information (PPI), and genomic data for at least one million US residents. AoURP EHR short-read whole genome sequence (srWGS) data was accessed via the version 7 curated data repository (CDR v7) within the Controlled Tier. Participants that were included in this cohort were recruited between 2018 and 2022. AoURP participants with both ABO blood types derived from serology and srWGS data were included in our analyses. Serology was obtained from the EHR domain “Labs & Measurements” under the logical observation identifiers names and codes (LOINC) code 882-1 (Abo and Rh group [Type] In Blood) and 883-9 (ABO group [Type] in Blood). For individuals with multiple ABO serology results, we excluded any individuals whose ABO blood type differed upon repeat testing.

We used computed ancestry provided by the AoURP consortium based on srWGS ^34,35^. The ancestry categories are based on those described in gnomAD^36^, Human Genome Diversity Project^37^, and 1KGP^27^: African (AFR), Latino/Native American/Admixed American (AMR), East Asian (EAS), European (EUR), Middle Eastern (MID), and South Asian (SAS). A random forest classifier was trained on a set of the HGDP and 1000 Genomes samples, and 16 principal components (PCs) of the training sample genotypes were generated. AoURP samples were projected into the PC analysis space of the training data, and the classifier was applied to determine ancestry.

### Derivation of ABO Blood Type and Alleles in the AoURP

srWGS was accessed in the Hail MatrixTable format from the v7 genomics CDR. We extracted SNPs of interest, including tSNPs utilized in existing studies and functional ABO variants from the AoURP curated subset files “srWGS: ACAF Threshold” that is comprised of srWGS SNP and insertion-deletion (indel) variants that are frequent in the AoURP computed ancestry subpopulations. Using SHAPEIT5^38^, we phased a ∼1 million base pair locus that encompassed the *ABO* gene with the 1KGP phase three collection used as the reference.

ABO alleles were derived using functional variants and then tSNPs. For deriving ABO alleles from functional variants, we utilized a 2-SNP approach. Individuals were first assessed on whether they had O or non-O blood type alleles using the functional variant rs8176719 with T engendering O alleles and TC engendering non-O alleles. Those with non-O alleles were further assessed to determine if they had A or B alleles using the functional variant rs8176746 with G engendering A alleles and T engendering B alleles. Blood types were inferred based on whether an individual had the following alleles: O/O (O), A/O or A/A (A), B/O or B/B (B), A/B (AB).

We then derived ABO alleles using tSNPs from existing studies. We first derived only O vs non-O alleles using the O vs non-O tSNPs: rs505922 (C>T), rs657152 (A>C), rs8176704 (G>A), rs687289 (A>G), rs612169 (G>A), rs529565 (C>T), rs8176693 (C>T), rs514659 (C>A), and rs8176645 (A>T). In each scenario the reference allele engendered the non-O allele, and the alternative allele engendered the O allele. Those with non-O/non-O or non-O/O alleles were considered to have a non-O blood type, and those with O/O alleles were considered to have an O blood type. To assess A vs B tSNPs, we utilized a 2-SNP approach. The functional variant rs8176719 was first used to determine O vs non-O, then A vs B alleles were derived using the A vs B tSNPs: rs8176749 (C>T), rs8176672 (C>T), rs8176741 (G>A), rs8176722 (C>A), rs8176720 (T>C). In each scenario the reference allele engendered the A allele and the alternative allele engendered the B allele. Blood types were subsequently determined using the alleles as previously described.

### Statistical Analysis for All of Us Research Program Cohort

LD between the functional variants and tSNPs was calculated using PLINK in the AoURP cohort overall (ALL) and in each ancestry-specific population (AMR, AFR, EUR, SAS, EAS, MID). Similarly, we calculated discordance between ABO blood types derived from functional variants versus tSNPs. Additionally, we calculated discordance between ABO blood types derived from SNPs (both functional variants and tSNPs), and those captured by serology. To evaluate the relationship between LD and discordance, we measured the relationship between the discordance observed between the O vs non-O functional variant and tSNPs, and the r^2^ calculated between the functional variant and tSNPs with Spearman’s correlation. Data from the *All of Us* Research Program is accessible only through the Researcher Workbench (https://workbench.researchallofus.org) as stipulated in the informed consent of participants in the program. This data use agreement prohibits investigators from providing row level data on AllofUs participants and thus providing a de-identified dataset is not possible for this manuscript. The code used for this demonstration project is available within the Researcher Workbench.

## Results

### Characteristics of the Studies for Systematic Review

The initial search retrieved 2,693 publications from seven databases and resulted in 136 publications that met criteria for inclusion in this review (**Figure 1**; **Table S6**). the mean and median year of publication were both 2016 **(Table 1**; **Figure S1**). Included studies were published in a variety of journals (**Table S7**) and focused on a wide variety of phenotypes with the majority of studies focused on hematological (n=28, 20.6%), cardiovascular (n=28, 20.6%), or oncology phenotypes (n=21, 15.4%) (**Figure S2**). The majority of platforms used fell under the genotyping category (n=72, 52.9%), followed by PCR/specific target (n=46, 33.8%), sequencing (n=5, 3.7%), and multiple platforms (n=13, 9.6%) (**Table 1**; **Figure S3**). A total of 56 (41.2%) of the studies did not describe use of a haplotype analysis, 52 (38.2%) did describe a haplotype analysis, and it was unclear for 28 (20.6%) of the studies (**Table 1**). Most studies (n=100, 73.5%) were done in populations that would be described as a single population group, AFR (n=7), AMR (n=3), EAS (n=30), EUR (n=59), and SAS (n=1). A total of 17 (12.5%) of the studies were done in multiple populations and 19 (14.0%) of the studies had an unclear population breakdown (**Table 1**; **Table S8**). Regardless of whether the study was done in a single or multiple populations, most studies were done in EUR populations (n=74, 56.9%).

**Table 1.**
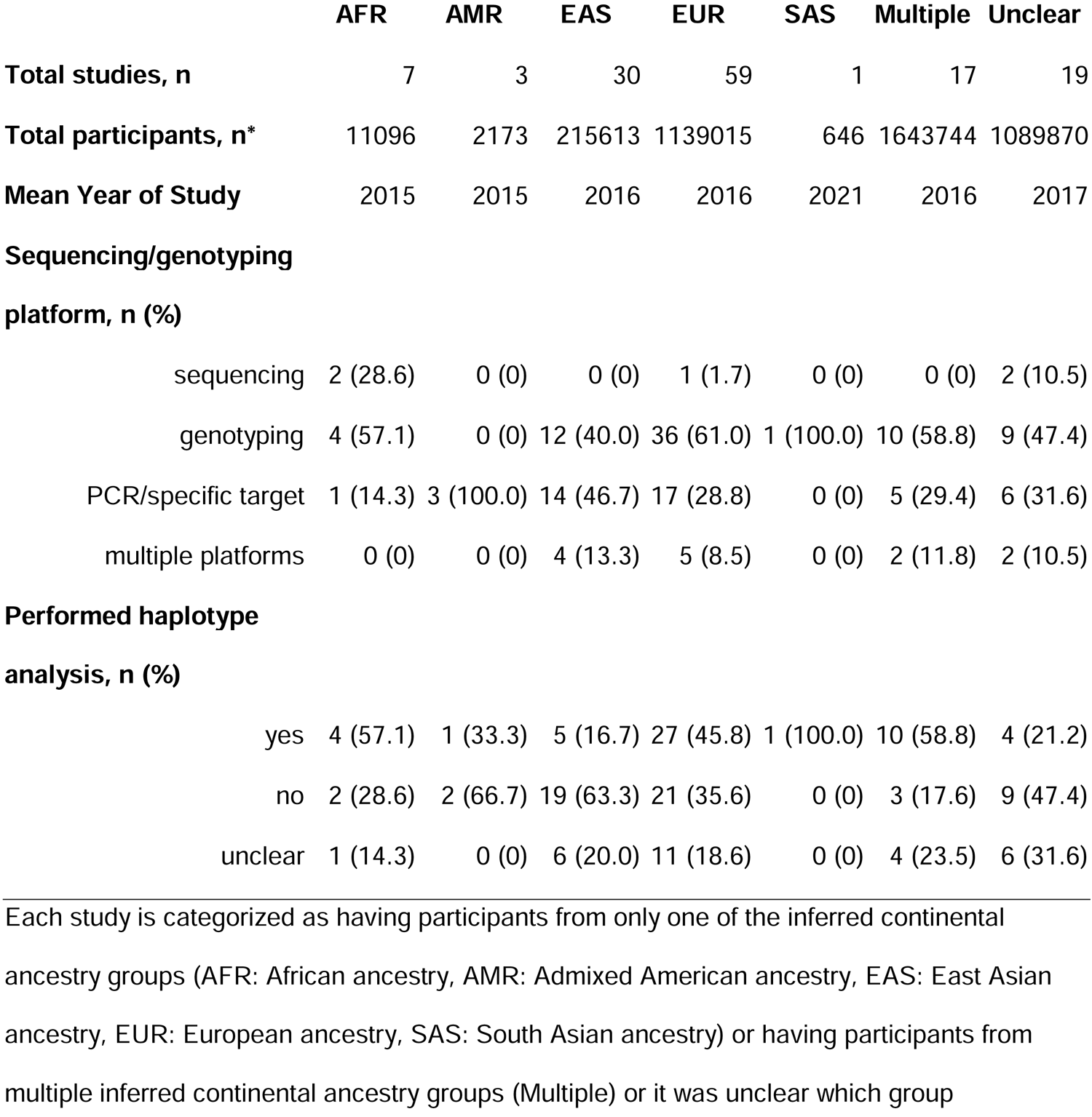
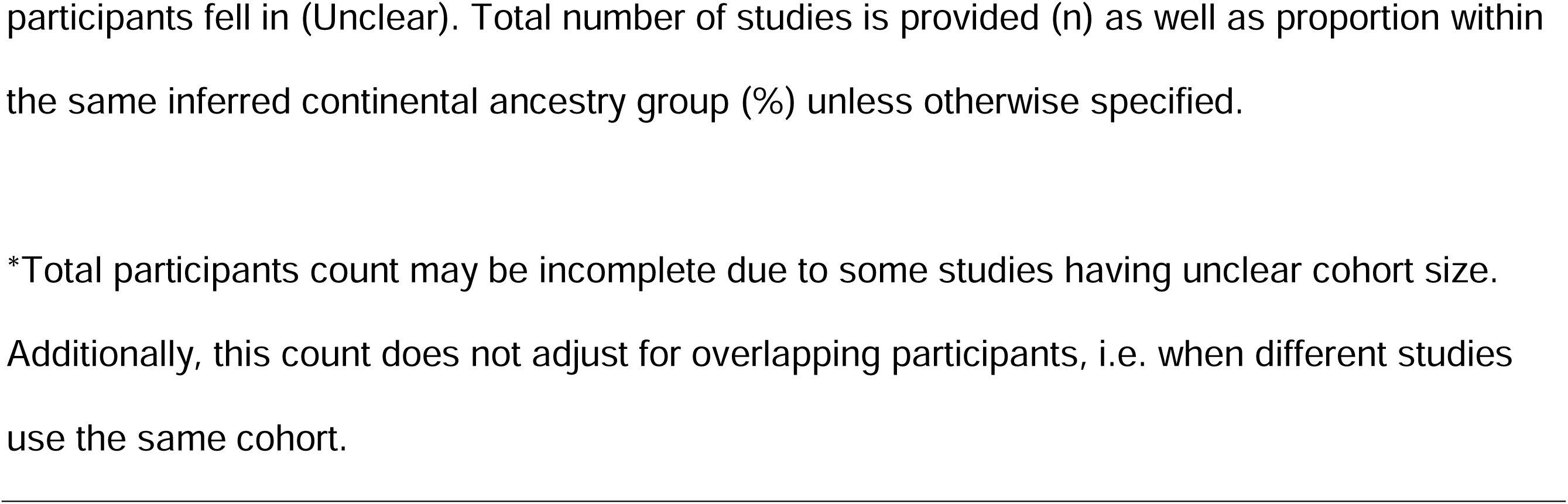
Summary of characteristics of systematic review studies.

|  | AFR | AMR | EAS | EUR | SAS | Multiple | Unclear |
| --- | --- | --- | --- | --- | --- | --- | --- |
| **Total studies, n** | 7 | 3 | 30 | 59 | 1 | 17 | 19 |
| **Total participants, n\*** | 11096 | 2173 | 215613 | 1139015 | 646 | 1643744 | 1089870 |
| **Mean Year of Study** | 2015 | 2015 | 2016 | 2016 | 2021 | 2016 | 2017 |
| **Sequencing/genotyping platform, n (%)** |  |  |  |  |  |  |  |
| sequencing | 2 (28.6) | 0 (0) | 0 (0) | 1 (1.7) | 0 (0) | 0 (0) | 2 (10.5) |
| genotyping | 4 (57.1) | 0 (0) | 12 (40.0) | 36 (61.0) | 1 (100.0) | 10 (58.8) | 9 (47.4) |
| PCR/specific target | 1 (14.3) | 3 (100.0) | 14 (46.7) | 17 (28.8) | 0 (0) | 5 (29.4) | 6 (31.6) |
| multiple platforms | 0 (0) | 0 (0) | 4 (13.3) | 5 (8.5) | 0 (0) | 2 (11.8) | 2 (10.5) |
| **Performed haplotype analysis, n (%)** |  |  |  |  |  |  |  |
| yes | 4 (57.1) | 1 (33.3) | 5 (16.7) | 27 (45.8) | 1 (100.0) | 10 (58.8) | 4 (21.2) |
| no | 2 (28.6) | 2 (66.7) | 19 (63.3) | 21 (35.6) | 0 (0) | 3 (17.6) | 9 (47.4) |
| unclear | 1 (14.3) | 0 (0) | 6 (20.0) | 11 (18.6) | 0 (0) | 4 (23.5) | 6 (31.6) |

Each study is categorized as having participants from only one of the inferred continental ancestry groups (AFR: African ancestry, AMR: Admixed American ancestry, EAS: East Asian ancestry, EUR: European ancestry, SAS: South Asian ancestry) or having participants from multiple inferred continental ancestry groups (Multiple) or it was unclear which group
participants fell in (Unclear). Total number of studies is provided (n) as well as proportion within the same inferred continental ancestry group (%) unless otherwise specified.
\*Total participants count may be incomplete due to some studies having unclear cohort size. Additionally, this count does not adjust for overlapping participants, i.e. when different studies use the same cohort.

A total of 11 SNPs were used to determine O vs non-O blood type alleles and a total of 9 SNPs were used to determine A vs B blood type alleles (**Figure S4**). Some studies used more than one SNP (n=9 [6.6%] to determine O vs non-O and n=20 [14.7%] to determine A vs B) while others used a single SNP (n=115 [84.6%] to determine O vs non-O and n=87 [64.0%] to determine A vs B) (**Table S9**). Out of the 136 studies, 67 (49.3%) used the functional variant (rs8176719) to determine the O blood type allele and 90 (66.2%) studies used at least one of the functional variants (rs7853989, rs8176743, rs8176746, rs8176747) to determine the A or B blood type allele.

### Linkage and Suitability Across Inferred Ancestry Groups for O vs non-O tSNPs Used by Studies in Systematic Review

tSNPs used to determine the O allele were in highest LD with the functional variant in the EAS population with median r^2^=0.95 (range 0.39-1) (**Figure 2**). No tSNP was in perfect LD with the O allele functional variant rs8176719 across all populations. In the AFR population, tSNPs used in prior studies displayed extremely low LD with the functional variant with median r^2^=0.36 (range 0.14-0.47) (**Figure 2B**). LDpop was unable to retrieve values for the tSNP rs72238104 used by one study in the EAS population^39^. We observed a wide range of LD between the reported tSNPs and the functional variant(s) across superpopulations, especially in the AFR group **(Figure 3)**. All tSNPs had significantly different medians across superpopulations (**Table S10**). Most notably, for seven out of the nine total tSNPs, the AFR population exhibited significantly lower r^2^ values when compared to EAS and SAS populations (**Table S11**).

**Figure 2.**
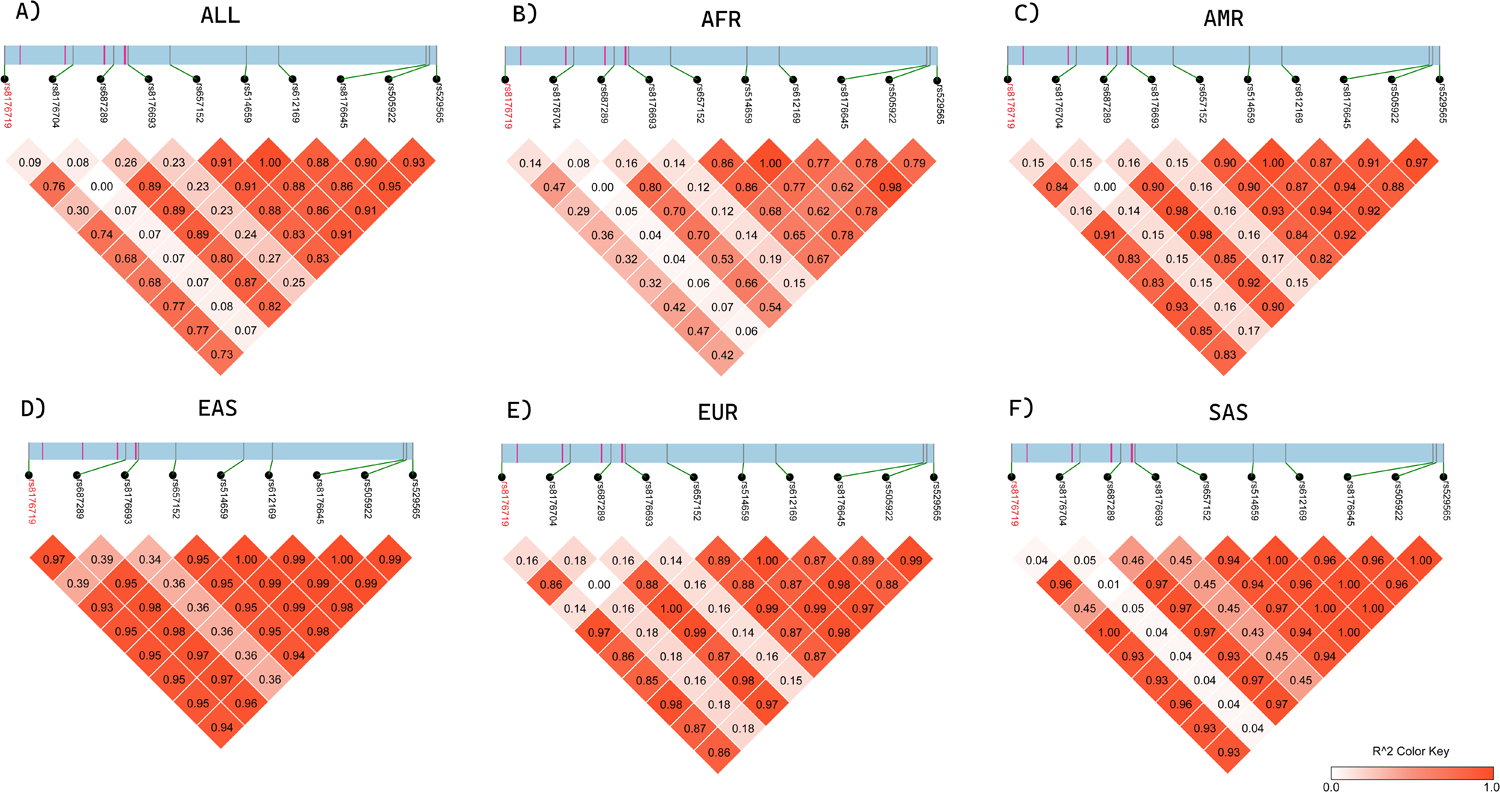
LD heatmap for SNPs that differentiate O vs non-O alleles across 1000 Genomes Project-defined superpopulations. This figure represents a 16.59 kb region of the ABO gene starting at 133.258 Mb and ending at 133.274 Mb. This region encompasses the locations of the O vs non-O functional variant and tag SNPs. The blue block represents the noncoding regions and the pink lines represent exons. The functional variant is highlighted in red. Linkage dissociation blocks representing the r^2^ values of the O vs non-O functional variant and tSNPs are shown beneath the section of the ABO gene. Lighter color blocks represent lower r^2^ values. **A)** Represents r^2^ values across all continental populations. **B) - F)** Represents r^2^ values across specific continental populations. AFR: African ancestry, AMR: Admixed American ancestry, EAS: East Asian ancestry, EUR: European ancestry, SAS: South Asian ancestry.

**Figure 3.**
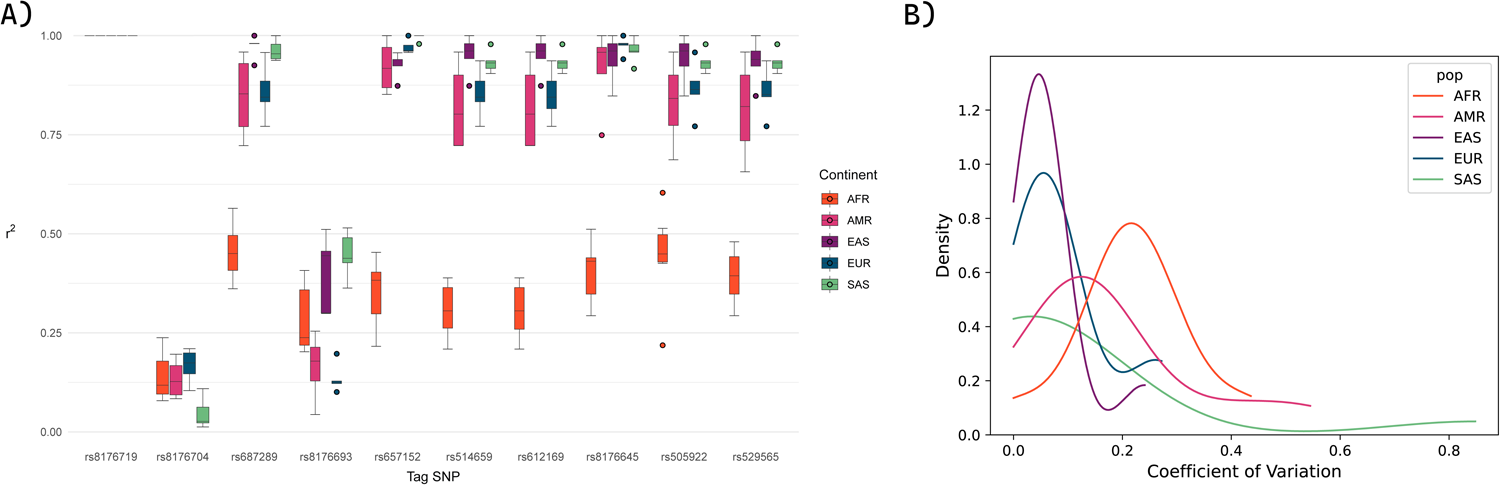
Linkage disequilibrium of O versus non-O tSNPs with functional variants across inferred continental ancestry groups. **A)** r^2^ between O vs non-O functional variant (rs8176719) and tag SNPs calculated across subpopulations and plotted per superpopulation (AFR: African ancestry, AMR: Admixed American ancestry, EAS: East Asian ancestry, EUR: European ancestry, SAS: South Asian ancestry). **B)** The coefficient of variation for r^2^ between the O vs non-O functional variant (rs8176719) and tSNPs calculated for each subpopulation population and plotted as a kernel density estimate plot across superpopulations.

In the primary analysis for the O vs non-O SNPs, when a study used a tSNP we compared r^2^ values for the reported tSNPs to the functional variant rs8176719 using inferred ancestry groups tSNPs had median r^2^=0.443 (range 0.387-0.448) in AFR, median r^2^=0.869 (range 0.853-0.975) in EUR, and median r^2^=0.946 (range 0.946-0.976) in EAS (**Figure 4A**). The single study that used a tSNP in an AMR population had r^2^ of 0.820. Tag SNPs used by studies had significantly lower r^2^ in AFR (median r^2^=0.443) than in EAS (median r^2^=0.946; p=1.1×10^-5^) and EUR (median r^2^=0.869; p=0.023). We also observed that tSNPs performed worse in EUR (median r^2^= 0.869) populations compared to EAS populations (median r^2^=0.946; p=1.6×10^-3^). Other pairwise comparisons of median r^2^ between populations were not statistically significant (**Table S11**).

**Figure 4.**
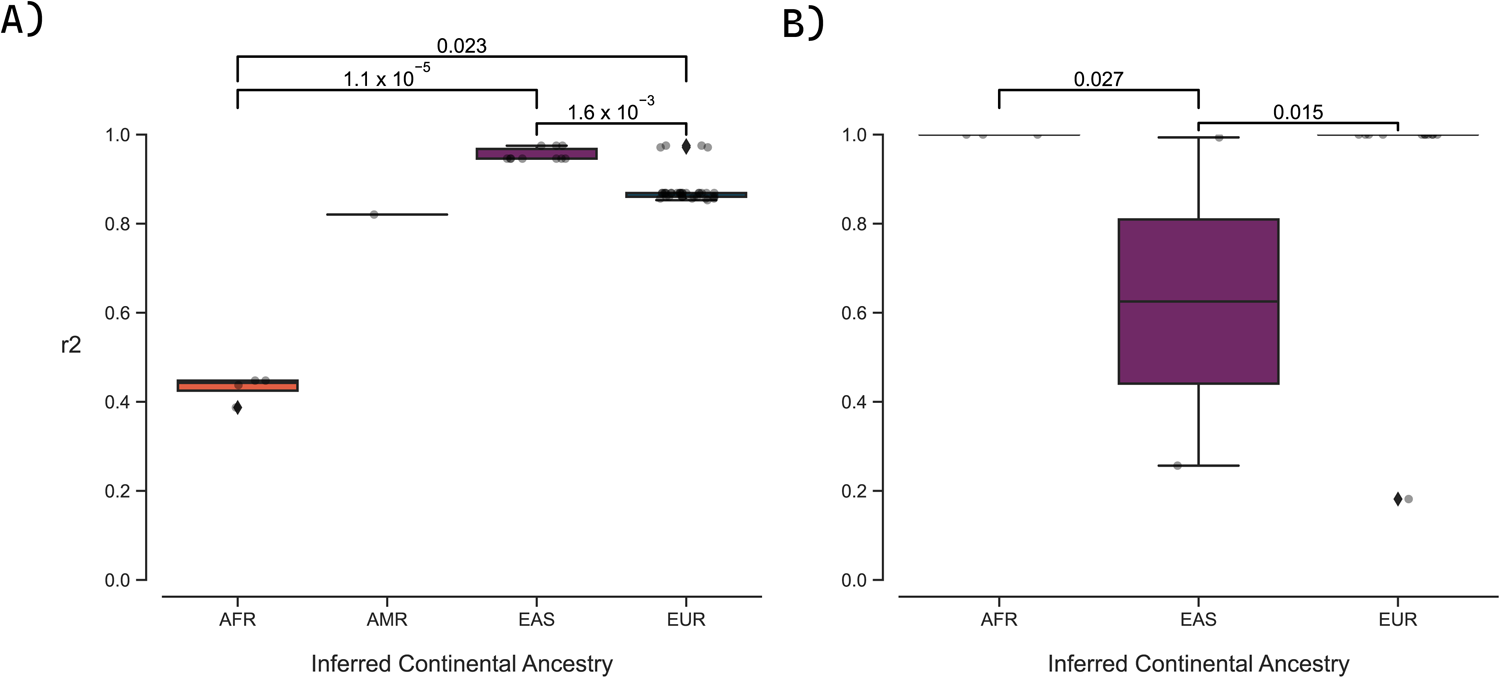
Linkage disequilibrium between tSNPs and functional variants for O vs. non-O (A) and A vs. B (B) by inferred continental ancestries for each study. r^2^ was calculated for each SNP used to determine ABO alleles for each study included in this systematic review that used a tSNP. r^2^ was calculated using the reference population (AFR: African ancestry, AMR: Admixed American ancestry, EAS: East Asian ancestry, EUR: European ancestry) that best matched the populations used by each study. If a study used more than one population, then that study is included more than once as each population may have a different r^2^ value. For studies using more than one SNP for a specific population, we assumed they were using the SNP with the highest r^2^ value for their population. **A)** rs8176719 was the functional variant used to calculate r^2^ which differentiates O vs non-O alleles and **B)** rs8176746 was selected as the representative functional variant which differentiates A vs B alleles and was used to calculate r^2^. No studies using an AMR population used a tSNP to derive A vs B blood type alleles.

### Linkage and suitability Across Inferred Ancestry Groups of A vs B tSNPs Used by Studies in Systematic Review

The four functional variants that differentiate between the A and B blood type alleles (rs7853989, rs8176743, rs8176746, rs8176747) were in very high LD with one another only in AFR, EAS, and SAS with range of r^2^=0.96-1 (**Figure S5**). In AMR and EUR, the functional variant rs7853989 had lower LD with the other three functional variants with r^2^=0.72 in AMR and r^2^=0.71 in EUR. In EAS, all the tSNPs were in high LD with one another with the exception of rs8176720, which was also in low LD with the other reported SNPs (r^2^ range 0.05-0.36 across all populations). Within and across superpopulations, we observed a wide range of LD between the reported tSNPs and the functional variant(s) (**Figures S5-S7**). Using rs8176746 as the A vs B representative functional variant, four (rs7853989, rs8176672, rs8176720, rs8176722) out of the seven tSNPs had significantly different medians across superpopulations (**Table S10**). Pairwise comparisons between superpopulations were further examined for these 4 tSNPS, but no one population consistently performed worse than the others (**Table S11**).

With respect to A vs B SNPs, the number of studies that used one of the functional variants (rs7853989, rs8176743, rs8176746, rs8176747) vs tSNPs were: AFR (78.6%; 21.4%), AMR (100%; 0%), EAS (92.9%; 7.1%), and EUR (75%, 25%). The two studies in AFR populations that used A vs B tSNPs both had r^2^=1 (**Figure 4B**). For studies in EAS populations, one study used a tSNP with r^2^= 0.9936 and one with a r^2^=0.2566. For studies using an EUR population, the r^2^ range was 0.1816-1 with a median of 1. No studies using an AMR population used a tSNP to infer A or B blood type alleles (**Figure 4B**). Pairwise comparisons of median r^2^ between populations were not statistically significant (**Table S12**).

### Linkage and Suitability of tSNPs from Studies in Systematic Review, Evaluated in the All of Us Research Program

A total of 10,771 individuals were identified with both srWGS and ABO blood type serology results in the AoURP, comprised of individuals with AFR (n=2,000; 18.6%), AMR (n=3,794; 35.2%), EAS (n=302; 2.8%), EUR (n=4,496; 41.7%), MID (n=47; 0.4%) and SAS (n=132; 1.2%) ancestry (**Table 2**). A majority of the cohort was female (n=7,774; 71.9%) and had a mean age of 53. Nearly half of the cohort had the O blood type (n=5,274; 49.0%) with the O blood type being most prevalent in individuals with AFR and AMR ancestry (n=1,004; 50.2% and n=2,179; 57.4%).

**Table 2.** Demographic characteristics for *All of Us* cohort.

|  | AFR | AMR | EAS | EUR | MID | SAS |
| --- | --- | --- | --- | --- | --- | --- |
| **Total, n** | 2000 | 3794 | 302 | 4496 | 47 | 132 |
| **Mean Age (Standard** |  |  |  |  |  |  |
| **Deviation)** | 50 (16.6) | 45 (16.1) | 53 (15.9) | 61 (16.8) | 50 (17.5) | 52 (16.2) |
| **Sex (female), n (%)** | 1535 (76.8) | 3066 (80.8) | 208 (68.9) | 2822 (62.8) | 30 (63.8) | 83 (62.9) |
| **Blood Type, n (%)** |  |  |  |  |  |  |
| O | 1004 (50.2) | 2179 (57.4) | 122 (40.4) | 1901 (42.3) | 18 (38.3) | 52 (39.4) |
| A | 553 (27.7) | 1104 (29.1) | 91 (30.1) | 1875 (41.7) | 22 (46.8) | 38 (28.8) |
| B | 384 (19.2) | 437 (11.5) | 73 (24.2) | 561 (12.5) | ≤20 (10.6) | 32 (24.2) |
| AB | 59 (3.0) | 74 (2.0) | ≤20 (5.3) | 159 (3.5) | ≤20 (4.3) | ≤20 (7.6) |

Each participant is estimated to have AFR (African), AMR (Admixed American), EAS (East Asian), EUR (European), MID (Middle Eastern), or SAS (South Asian) ancestry. Total number of participants is provided (n) as well as proportion within the same ancestry group (%) unless otherwise specified. Aggregate statistics corresponding to fewer than 20 participants are suppressed in compliance with AoURP data and statistics dissemination policy.

The median r^2^ for O vs non-O tSNPs (n=9) versus the functional variant was r^2^=0.75 (range 0.18-0.83) in the complete cohort (ALL) (**Figure S8).** Compared to the other ancestry groups, all the tSNPs were observed to have very low LD in the AFR ancestry group with median r^2^=0.44 (range 0.13-0.56). LD was consistently low across all ancestry groups for rs8176704 with median r^2^=0.12 (range 0.001-0.13) and rs8176693 with median r^2^=0.19 (range 0.14-0.39). The median r^2^ for A vs B tSNPs (n=5) versus the functional variant was r^2^=0.84 (range 0.15-0.998) in ALL. LD was consistently low across all ancestry groups for rs8176720 with median r^2^= 0.194 (range 0.076-0.305). Heatmaps of discordance between blood types derived from functional variants versus tSNPs are provided in the **Supplementary Figures S9** and **S10**.

### Performance of Genotype-Derived ABO Blood Types in the AoURP

We assessed discordance between O and non-O blood types derived from the functional variant rs8176719 and tSNPs, and those derived from serology in the complete cohort and across ancestry groups (**Figure 5A**). The tSNPs rs8176704 and rs8176693 displayed the most discordance across all ancestry groups with a median of 0.43 (range 0.38-0.58) and 0.46 (range 0.40-0.58) respectively. Compared to the other ancestry groups, discordance was particularly high across all SNPs in AFR with a median of 0.19 (range 0.01-0.51). In almost all cases, O and non-O blood types derived from the functional variant observed less discordance with serology than O and non-O blood types derived from tSNPs. In 10 cases, the tSNPs observed less discordance than the functional variant: rs505922 in EUR and SAS, rs687289 in EUR and SAS, rs612169 in EUR and SAS, rs529565 in EUR and SAS, and rs514659 in EUR and SAS. The majority of the discordance observed in these case occurred in the same 120 EUR participants and ≤20 EAS participants across the tSNPs. Additionally, LD (r^2^) significantly correlated with discordance with Spearman’s ρ=-0.90 and p-value=3.08×10^-23^ (**Figure 5B**).

**Figure 5.**
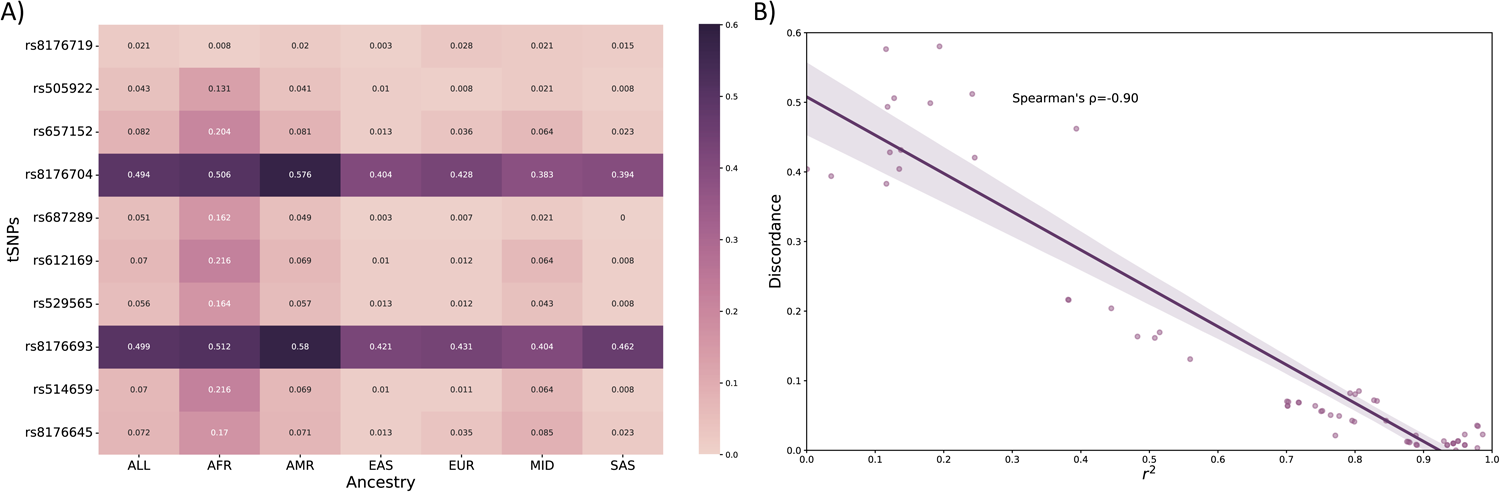
Discordance between O vs non-O blood types from tSNPs and serology. **A)** Discordance between ABO O and non-O blood types derived from the O vs non-O functional variant (rs8176719) and tSNPs and those derived from serology are displayed as a heatmap. Discordance (proportion of non-matches in a cohort of 10,771) was calculated across all ancestry groups (ALL) and calculated for each inferred continental ancestry group separately (AFR: African ancestry, AMR: Admixed American ancestry, EAS: East Asian ancestry, EUR: European ancestry, MID: Middle Eastern ancestry, SAS: South Asian ancestry). **B)** Discordance between blood types derived from O vs non-O tSNPs and those derived from serology was plotted along with LD (r^2^) between the O vs non-O functional variant and tSNPs. A Spearman’s correlation coefficient was calculated (−0.90, p-value = 1.51 x 10^-38^).

## Discussion

In the absence of serology, ABO blood types and alleles are often determined using genomic variants and frequently derived in the pursuit of investigating associations or risk with cardiovascular disease^9,40–42^. Here we present the results of a systematic review and analysis that aimed to understand if studies are using appropriate tSNPs for their population to determine ABO alleles, and if tSNPs are transferable across different populations. We found that many tSNPs used to differentiate between O vs non-O alleles had low r^2^ with the functional variant, particularly for underrepresented populations such as AFR and AMR populations. This non-transferability of O vs non-O tSNPs was further exemplified by the disparate LD structures we observed in the ABO gene across continental populations. Not only did we observe highly variable LD structure for reported SNPs across continental populations, but we also observed a wide range of linkage between the reported tSNPs and the functional variant(s) within populations. Furthermore, in a large and diverse cohort from AoURP, we found that O and non-O blood types derived from tSNPs had generally high discordance with serology, particularly in the population with African ancestry. The observed discordance was highly inversely correlated with LD.

The lack of diversity in genetic research has been well cited^2,28,43–45^, and the issue is compounded when findings done in one population are erroneously assumed to translate directly to another population^46–48^. Genetic studies have been disproportionately performed in cohorts of European ancestry with low representation of AFR and AMR individuals^43,46^ and our systematic review reflects this trend. Despite the research bias towards European populations, our study shows that ABO tSNPs generally performed the best in EAS populations. This likely reflects the early ABO genetic research performed primarily in Japanese cohorts^14,49^. Interestingly, we observed reductions in LD and blood group accuracy for tSNPs in EUR compared to EAS populations. Our results suggest that many of the tSNPs that were originally identified in EAS populations by early ABO researchers have been used without appropriate vetting in more diverse populations.

Acknowledging that tSNPs are not always transferable across populations also has broad implications for genomic research, which claims to be making efforts towards increasing cohort diversity^50^. While many studies established a tSNP linkage threshold of r^2^≥0.8^4,51,52^, often for genotyping and array purposes^53–55^, other studies acknowledge that the most conservative approach to selecting tSNPs is to select tSNPs that are a perfect proxy (r^2^=1.0) for the SNP of interest^52^. Additionally, discordance of ABO blood alleles derived from functional variants and tSNPs increases linearly as LD decreases in a diverse population, with a clear delineation observed at r^2^=0.9. Our results suggest that a conservative and very high linkage (r^2^>0.9) threshold is needed to maintain blood type accuracy in the population of interest. As seen in this study, tSNPs used for inferring O vs non-O alleles are not transferable across populations particularly in AFR and AMR populations. Even within superpopulations (AFR, AMR, EAS, EUR), tSNPs exhibit wildly varied linkage. A more rigorous approach would be to forgo the use of tSNPs and instead use functional variants, although we recognize that many genotyping methods cannot capture indels, such as the functional variant that differentiates between O vs non-O blood type alleles (rs8176719). Imputation of indels may be unreliable in some populations since accuracy is dependent on the matching of LD structure between a cohort of interest and a reference population^56^. In many cases, no appropriate reference population or very small reference populations are available for underrepresented continental ancestry groups, and this may lead to reduced imputation accuracy ^57–59^. Furthermore, many programs for phasing and imputation are not able to handle indels ^60,61^.

To evaluate the real-world consequences of utilizing tSNPs in place of functional variants, we utilized a large and diverse subset of the AoURP ^28^ cohort to observe discordance between ABO blood types derived from functional variants, tSNPs, and serology. Discordance was generally high between O and non-O blood types derived from the functional variant and those derived from serology in participants with AFR ancestry. This finding was unsurprising as LD calculated between the functional variant and tSNPs in AFR populations from 1KGP and AoURP were consistently low. Using tSNPs to derive ABO blood groups would have an increased likelihood of being inaccurate in under-represented populations such as those with African ancestry. Our data indicates that, depending on the tSNP used to derive O vs non-O blood types and the population of interest, as many as 58 out of a 100 people could be mistyped. Previously observed associations in ABO and the corresponding conclusions based on these potentially inaccurate blood types are likely to further exacerbate the already prevalent health disparities in genomic research.

We also observed inconsistent and inadequate documentation provided by publications. Very few studies described their population in terms of continental genetic ancestry and instead used only racial or ethnic terms which makes evaluating the appropriateness of a tSNP challenging. For a large portion of the studies, it was unclear whether the study conducted a haplotype analysis. This is relevant because, to determine ABO alleles, phased data and a haplotype analysis must be performed. Alarmingly, we were able to confirm that only 38.2% of the studies conducted a phasing or haplotype analysis, the lack of which would likely result in reduced accuracy of ABO blood group assignment.

This study has several limitations worthy of mention. Many studies only described their populations using racial or ethnic terms, and we were forced to infer continental genetic ancestry based off this poor proxy. We acknowledge the limitations of this approach and that, while categories such as race and ethnicity have historically been used as proxies for genetic ancestry due to positive correlation, race and ethnicity are socially-constructed concepts that should not be conflated with the biologically based groupings derived from genetic ancestry^62^. Similarly, even the populations described by the most diverse publicly available dataset, the 1KGP^27^, did not describe an appropriate continental ancestry population associated with Middle Eastern individuals. This reflects a larger issue that much work is needed to establish reference files that are more comprehensive and global. Secondly, some studies used more than one SNP to determine O vs non-O or A vs B blood type alleles but did not describe in which scenario each SNP was used. Thus, we were compelled to select the best performing SNP and our resulting analyses may reflect more optimistic results than the reality. In the systematic review portion of this study, we were not able to identify individuals that overlapped in multiple studies and adjust this total count in our analyses. Lastly, we were unable to account for complex haplotypes, rare O-determining variants, or non-ABO variation conferring changes in blood type serology.^63–65^. For instance, individuals of the Bombay and para-Bombay phenotypes appear to have functional A and B ABO alleles, but lack functional alleles in FUT1 and FUT2, which encode precursor substrates for A and B transferases. Consequently, we are unable to evaluate whether the few instances where blood types derived from tSNPs displayed less discordance with serology than with blood types derived from the functional variant was due to inaccuracies from serological typing, problems with sequence accuracy known to be associated with indels^66,67^, or rare functional variation in ABO or related genes.

## Conclusion

ABO blood types have widespread clinical use and are robustly associated with cardiovascular disease. Many studies opted to use functional variants to determine ABO blood alleles, however, as observed in this study, there is potential for misinference of ABO blood types due to using inappropriate tSNPs thus potentially leading to erroneous conclusions. We observed that LD structure among tSNPs used in studies to infer ABO alleles varied substantially across populations. Even within superpopulations (AFR, AMR, EAS, EUR, SAS), linkage between a tSNP and a functional variant varied widely. Many studies that used tSNPs for determining O vs non-O alleles that had low r^2^ particularly in AFR and AMR populations. Furthermore, high discordance was observed between O and non-O blood types derived from tSNPs versus serology, particularly in the AFR population from AoURP. Our results not only have important implications for how ABO alleles are inferred from genomic data, but also highlight that tSNPs are not always transferable across continental populations and may result in furthering the genomic research and health disparities^45^ already seen in underrepresented populations.

## Supporting information

ABO Supplementary Materials

## Availability of data and materials

Data from the *All of Us* Research Program is accessible only through the Researcher Workbench (https://workbench.researchallofus.org/workspaces/aou-rw-a90a6fe4/duplicateofabophewas) as stipulated in the informed consent of participants in the program. This data use agreement prohibits investigators from providing row level data on *All of Us* participants and thus providing a de-identified dataset is not possible for this manuscript. The code used for this demonstration project is available within the Researcher Workbench. Datasets not generated from the *All of Us* Research Program and the associated code are available on GitHub (https://github.com/Karnes-Lab/ABO_systematic_review).

## Acknowledgements

The authors would like to thank the participants of the *All of Us* Research Program. Research reported in this publication was supported by the National Institutes of Health’s National Heart, Lung, and Blood Institute (grants R01 HL156993, R01 HL158686, R21HL172036) and the Office of the Director (grant OT2OD036485).

## Author contributions

Conceived the study, J.H.K., K.L.M., J.B.G, J.R.M.; databases searches for systematic review, J.R.M., K.L.M.; data curation for systematic review, K.L.M., C.U.S., M.L.B, K.B., G.Q., J.F.; data analysis, K.L.M., A.K.; analysis support, C.U.S.; visualization, K.L.M., A.K., wrote the paper, K.L.M, J.H.K., A.K., with input from all authors.

