## Supplementary material for "Disparities in ABO Blood Type Determination Across Diverse Ancestries: A Systematic Review and Validation in the *All of Us* Research Program": ABO Supplementary Materials

*Correspondence to:

Jason H. Karnes, PharmD, PhD

Associate Professor

**SUPPLEMENTARY METHODS**

**ABO Blood Type**

The *ABO* gene in humans is responsible for encoding the enzymes that result in blood types, A, B, AB, and O. The ABO gene encodes glycosyltransferases that are required to change the existing antigens on red blood cells (RBCs) to A and/or B antigens. The O blood type arises when no glycosyltransferases are encoded by the ABO gene^1^. When inferring ABO blood types from genomic data, the most frequent functional variant that differentiates O vs non-O blood type alleles is rs8176719. Four functional variants (rs7853989, rs8176743, rs81767846, and rs8176747)^2^ differentiate A vs B blood type alleles although it is common to use a single functional variant instead of all 4 functional variants. Thus, a haplotype approach must be taken when determining ABO alleles as represented in **Supplementary Table S5**. Once ABO alleles are determined then an indivdidual’s ABO blood type can be determined. Individuals with O/O have an O blood type, those with A/O or A/A have an A blood type, those with B/O or B/B have B blood type, and those with both an A and B blood type allele have AB blood type.

**Search Strategy**

PICO Criteria

We conducted a systematic search for recent peer-reviewed publications that used SNPs to determine the primary ABO alleles (A, B, and O). We used the PICO format (Population, Intervention/exposure, Comparison and Outcome)^3^ (**Supplementary Table S1**) to help us define and inform our search strategy as.

Search Queries

The first time a study used single SNPs to determine ABO blood type from genetic data was in 2005,^4^ therefore we started our study search for this review in the year 2005. The following bibliographic databases were searched: PubMed/Medline (National Library of Medicine, 1946-2022), Embase (Elsevier, 1947-2022), Scopus (Elsevier, 1788-2022), Ovid/Medline (Wolters Kluwer, 1946-2022), and CINHAL Plus with Full Text (Ebsco, 1937-2022). In addition, Dissertations and Theses Global (ProQuest, 1861-2022) was also searched. The main keywords were: ("ABO Blood-Group System"[MESH]) OR ("Blood Group System*" AND (ABO OR ABH OR H OR "H type 1 Antigen")) OR ("Blood-Group System*" AND (ABO OR ABH OR H OR "H type 1 Antigen")) OR ("Blood Factor*" AND (ABO[ALL] OR ABH OR H OR "H type 1 Antigen")) OR ("Blood Type A") OR ("Blood Type" AND (B OR O OR AB)) OR "ABO Factor*" OR (“ABO Blood Group Antigen*”). These searches were conducted within 1 week of August 2022. The complete queries conducted in each database are provided in **Supplementary Table S2**.

Inclusion and Exclusion Criteria and Data Extraction

We used Endnote 20 (Clarivate, Philadelphia, PA, USA) as our literature reviewer software to compile our studies conducted in our initial search. During this process we removed initial duplicates automatically using the Endnote 20 software. We then uploaded these studies, as well as those identified using Google Scholar, to DistillerSR (DistillerSR, Ottawa, Canada) and proceeded to use this as our primary literature reviewer software that enabled us to efficiently screen references and extract data. Using DistillerSR, we were able to create screening and data extraction forms.

The publications meeting the following criteria were included for the initial title and abstract evaluation: 1) available in the English language, 2) the organism being studied is human, and 3) is a primary article. Other publications such as comments, editorials, review articles and conferences proceedings and abstracts were not included. The data dictionary for the extraction form is as described in **Supplementary Table S3**.

A secondary evaluation was carried out where we screened the full text and included only peer-reviewed articles that used single SNPs to derive ABO alleles (A, B, and O) from genomic data. During this step we also excluded clinical trials and studies that were using extensive haplotype methods to derive ABO blood type or subtype alleles since our primary question focuses on the use of tag SNPs (tSNPs) to derive ABO alleles from genomic data. The secondary screening question was included in the data extraction form that is described in **Supplementary Table S4**.

The bibliographic information associated with each reference was automatically extraction by the DistillerSR software. The primary data were extracted by two investigators independently using the following data extraction form described in **Supplementary Table S6**.

**Assessment of ABO Genotyping Platforms**

Because genotyping technology influences the functional ABO variation that could be determined, we also collected sequencing and genotyping platforms used to determine the SNPs and ABO alleles. We categorized these platforms into four groups: 1) sequencing, 2) genotyping, 3) PCR/specific target, and 4) multiple genotyping/sequencing platforms. The sequencing category describes a platform that results in information of a complete genome or exome. Genotyping describes platforms that do not sequence the whole genome but instead provides information on hundreds of thousands of DNA variants at specific locations in the genome and includes data derived from microarray approaches. PCR/specific target describes platforms that genotype individuals for a small number of DNA variants.

**Inferred Continental Ancestry**

We inferred genetic continental ancestry as described by the International Genome Sample Resource (IGSR) for their 1000 Genomes Project19 phase three collection^5^. The 1000 Genomes Project ran between 2008 and 2015 and created a large and diverse public catalogue of genotype data from 2,504 individuals from 26 populations in Africa (AFR), the Americas (AMR), East Asia (EAS), Europe (EUR), and South Asia (SAS). However, the studies collected for this systematic review did not consistently describe the genetic continental ancestry of the individuals that participated in their studies. For example, many studies described their populations using racial terms such as “White” or “Caucasian” or in terms of the population’s geography such as being “Japanese” or “Finnish”. Thus, in scenarios where continental ancestry was not provided, we inferred continental ancestry based on these other descriptors and categorized them into AFR, AMR, EAS, EUR, or SAS. For example, populations described as “White” or “Caucasian” would be labeled as EUR, while those described as being “Japanese” would be labeled as EAS. One study was difficult to approximate as it described a population recruited across the United Arab Emirates. As there is no specific Middle Eastern ancestral group described by the 1000 Genomes Project, we categorized this population into the geographically nearest 1000 Genomes Project sub-population which is the Punjabi (PJL) population from Lahore, Pakistan and is part of the SAS continental population. We acknowledge the limitations of this approach and that, while categories such as race and ethnicity have historically been used as proxies for genetic ancestry due to positive correlation, race and ethnicity are socially constructed concepts that should not be conflated with the biologically based groupings derived from genetic ancestry.

**Extracted & Derived Data**

The extracted and derived data used for this study are available as a separate CSV document (**Supplementary Table S6)**. Extracted data includes bibliographic information automatically extracted by DistillerSR, and data extracted by reviewers as described in **Supplementary Table S4**. Derived data includes: inferred continental ancestry, sequencing/genotyping category, and phenotype category.

**Statistical Analysis**

We visually examined LD structure of the reported SNPs across populations using LDBlockShow^6^ and data from the 1000 Genomes Project on GRCh38^5^. Reported SNPs for O vs non-O alleles were the following: rs8176719 (functional variant), rs8176704, rs687289, rs8176693, rs657152, rs514659, rs612169, rs8176645, rs505922, rs529565, and rs72238104. However, the SNP rs72238104 was not able to be found in the NIH Single Nucleotide Polymorphism Database (dbSNP). Reported SNPs for A vs B alleles were the following: rs8176749, rs8176747 (functional variant), rs8176746 (functional variant), rs8176743 (functional variant), rs8176741, rs7853989 (functional variant), rs8176722, rs8176720, and rs8176672. r^2^ was calculated between functional variants and all reported tSNPs for each superpopulation described in the 1000 Genomes Project (AFR, AMR, EAS, EUR, and SAS) except for the reported SNP rs72238104. These results are presented in **Figure 2** and **Supplemental Figure S5**.

To examine within and across population variation of linkage, we calculated r^2^ between all reported tSNPs and functional variants across the 26 subpopulations described in the 1000 Genomes Project. The subpopulations that make up the African (AFR) superpopulation are the following: Yoruba in Ibadan, Nigera (YRI); Luhya in Webuye, Kenya (LWK); Gambian in Western Gambia (GWD); Mende in Sierra Leone (MSL); Esan in Nigera (ESN); Americans of African Ancestry in SW USA (ASW); African Caribbeans in Barbados (ACB). The subpopulations that make up the Admixed American (AMR) superpopulation are the following: Mexican Ancestry from Los Angeles, USA (MXL); Puerto Ricans from Puerto Rico (PUR); Colombians from Medellin, Colombia (CLM); Peruvians from Lima, Peru (PEL). The subpopulations that make up the East Asian (EAS) superpopulation are the following: Han Chinese in Bejing, China (CHB); Japanese in Tokyo, Japan (JPT); Southern Han Chinese (CHS); Chinese Dai in Xishuangbanna, China (CDX); Kinh in Ho Chi Minh City, Vietnam (KHV). The subpopulations that make up the European (EUR) superpopulation are the following: Utah Residents from North and West Europe (CEU); Toscani in Italia (TSI); Finnish in Finland (FIN); British in England and Scotland (GBR); Iberian population in Spain (IBS). The subpopulations that make up the South Asian (SAS) superpopulation are the following: Gujarati Indian from Houston, Texas (GIH); Punjabi from Lahore, Pakistan (PJL); Bengali from Bangladesh (BEB); Sri Lankan Tamil from the UK (STU); Indian Telugu from the UK (ITU). Values were then grouped by superpopulation to descriptively evaluate variation within and across superpopulations. The functional variant for O vs non-O alleles was defined as rs8176719 and the functional variants for A vs B alleles were defined as rs7853989, rs8176743, rs8176746, and rs8176747. For our primary analyses the functional variant rs8176746 was used as the representative functional variant for A vs B alleles to calculate r^2^ values. We first conducted a Kruskal Wallis test identify differences in medians across superpopulations for each tSNP (**Supplementary Table S10**). Then we did pairwise comparisons across superpopulations within each tSNP using Dunn’s test of multiple comparisons(**Supplementary Table S11**). Summary statistics are provided as a separate CSV document (**Supplementary Table S15**). We also calculated the coefficient of variation for r^2^ in each 1000 Genomes Project superpopulation to descriptively evaluate variance across each superpopulation in a kernel density plot made using Python. These results are presented in **Figure 3** and **Supplemental Figures S6** and **S7**.

In our primary analysis, we first inferred the continental ancestry of the population(s) used in each study. However, for all subsequent analyses, we excluded a single study that had a population with inferred South Asian (SAS) continental ancestry due to the small sample size, restricting our inferred continental ancestries of interest to AFR, AMR, EAS and EUR. We then calculated a single r^2^ value for each population in each study between the study’s reported tSNP and the functional variant (rs8176719 for O vs non-O and rs8176746 for A vs B) using the matching 1000 Genomes Project superpopulation. We compared medians of r^2^ values between superpopulations with Dunn’s test of multiple comparisons using rank sums after doing a Kruskal Wallis test using R^7^ (**Supplementary Table S12**). Analyses were performed separately for O vs non-O tSNPs and A vs B tSNPs. Our primary analysis excluded studies that used ABO allele-determining functional variants, however a secondary analysis was also performed that included studies using functional variants (**Supplementary Figure S11**).

Additionally, we calculated the proportion of individuals (**Supplementary Figure S12**) as well as the proportion of studies (**Supplementary Figure S13**) whose ABO alleles were potentially being mis-inferred due to poor performing tSNPs, defining poor proxy SNPs as SNPs with r^2^<0.9. We compared these proportions across inferred ancestry groups using Fisher’s Exact Test adjusted for pairwise comparisons in R^7^ (**Supplementary Tables S13** and **S14**).

Lastly, we sought to evaluate the real-world consequences of utilizing tSNPs in place of functional variants in a large and diverse cohort of 10,771 participants. We utilized a subset of the *All of Us* cohort that encompassed participants that had available short-read whole-genome sequenced (srWGS) data and data from electronic health records (EHR) that described ABO blood types derived from serology (**Table 2**). LD between the functional variants and tSNPs was assessed in the *All of Us* cohort. r^2^ was calculated between the functional variants and tSNPs using PLINK in the complete cohort and in each ancestry group (**Supplementary Figure S8**).

We calculated discordance (the proportion of non-matching pairs) between ABO blood types derived from functional variants and tSNPs in the complete cohort (ALL), and in each ancestry-specific population (AMR, AFR, EUR, SAS, EAS, MID). Although not a superpopulation represented in the 1000 Genomes Project superpopulations offered in LDLink, we included those that had Middle Eastern (MID) global ancestry predictions. We calculated discordance between ABO O and non-O blood types derived from the O vs non-O functional variant (rs8176719) and those derived from tSNPs that differentiate O vs non-O blood types (rs505922, rs657152, rs8176704, rs687289, rs612169, rs529565, rs8176693, rs514659, rs8176645) (**Supplementary Figure S9**). We also assessed ABO blood types derived from A vs B tSNPs. ABO blood types were also derived with the two functional variants rs8176719, which differentiates between O vs non-O alleles, and rs8176746, which differentiates between A vs B alleles. These blood types were then compared to ABO blood types derived from the functional variants that differentiate O vs non-O alleles (rs8176719) and tSNPs that differentiate between A vs B alleles (rs8176749, rs8176672, rs8176741, rs8176722, rs8176720) (**Supplementary Figure S10B**).

Additionally, we calculated discordance between ABO blood types derived from SNPs (both functional variants and tSNPs), and those captured by serology. LD, in the form of r^2^, was calculated in this cohort as well as the 1000 Genomes Project cohort between the functional variants and tSNPs using PLINK in the complete cohort and in each ancestry group (AMR, AFR, EUR, SAS, EAS, MID) (**Figure 5A**; **Supplementary Figure S10A**).

We measured the relationship between the discordance observed between O and non-O blood types derived from O vs non-O tSNPs and those derived from functional variants, and LD (r^2^) between the O and non-O tSNPs and functional variant with a Spearman’s correlation (**Figure 9B**). We also measured the relationship between the discordance observed between O and non-O blood types derived from O vs non-O tSNPs and those derived from serology, and LD (r^2^) between the O and non-O tSNPs and functional variant with Spearman’s correlation (**Figure 5B**).

*Linkage and suitability of tSNPs used by Systematic Review studies across inferred ancestry groups*

For O vs non-O, tSNPs that were considered poor proxies (r^2^<0.9) for the functional variant were used for all AFR individuals (n=6,234) and all AMR individuals (n=2,077). All EAS individuals (n=133,293) were genotyped with tSNPs having r^2^≥0.9 and 280,259 (42.9%) of EUR individuals were genotyped with tSNPs having r^2^<0.9. Proportions of tSNPs with r^2^<0.90 were significantly different for each pairwise comparison between populations (p=0 for all pairwise comparisons with the exception of AFR versus AMR) (**Supplementary Figure S12A**; **Supplementary Table S13**). When comparing across studies, the number of studies that used tSNPs with r^2^<0.9 were only significantly different between AFR (n=4; 100%) and EAS (n=0; 0%; p=0.005), and between EAS and EUR (n=33; 89%) with p=1.6x10^-6^ (**Supplementary Figure S13A**; **Supplementary Table S14**).

For A vs B, tSNPs for all AFR individuals (n=6,151) had r^2^≥0.9. For EAS, 41.6% (n=92) had r^2^<0.9 and 58.4% (n=129) had r^2^≥0.9. For EUR, 6.4% (n=4,327) had r^2^<0.9 and 93.6% (n=63,594) had r^2^≥0.9. These proportions were also significantly different for each pairwise comparison: (p=1.99x10^-144^ for AFR vs EAS, p=1.35x10^-168^ for AFR vs EUR, p=3.79x10^-50^ for EAS vs EUR) (**Supplementary Figure S12B**; **Supplementary Table S13**). When comparing across studies, the number of studies that used tSNPs with r^2^<0.9 where not statistically significant between inferred continental ancestry groups (**Supplemental Figure S13B**; **Supplementary Table S14**).

*Performance of genomic tSNP-derived blood types in the All of Us Cohort*

We assessed discordance between O and non-O blood types derived from the functional variant rs8176719 and those derived from tSNPs in the complete cohort and across estimated global ancestry groups (**Supplementary Figure S9A**). The tSNPs rs8176704 and rs8176693 displayed the most discordance across all ancestry groups with a range of 0.362-0.557 and 0.383-0.561 respectively. Across most tSNPs, the AFR ancestry group showed the most discordance (range 0.136-0.504) and the EAS (0.007-0.417) and SAS (range 0.008-0.447) ancestry groups showed the least discordance. We also assessed the relationship between discordance and LD (**Supplementary Figure S9B**). LD (r^2^) significantly correlated with discordance (Spearman’s ρ=-0.97 and p-value=1.51 x 10^-38^). Discordance was also assessed between ABO blood types derived from the two functional variants and from blood types derived from the O vs non-O functional variants and the A vs B tSNPs (**Supplementary Figure S10B**). There was virtually no discordance observed for the tSNP rs8176672 across all populations. Across all ancestry groups and tSNPs the discordant range was 0-0.05.

**Data and Code Availability**

Python and R scripts related to analyses performed in association with the systematic review are made available on GitHub (*TBA*). The extracted and derived data used for this study are available as a separate CSV document (**Supplementary Table S6)**. Scripts and data related to analyses performed in the *All of Us* cohort are available on the Research Workbench (*TBA*).

**SUPPLEMENTARY TABLES**

**Supplementary Table S1.** PICO criteria for research question.

| Parameters | Description |
| --- | --- |
| Population | genetic study published between the years 2005-2022 that is defining its participants’ ABO blood group alleles with SNPs – either tag SNPs or functional variant |
| Intervention/exposure | tag SNP |
| Comparison/control | functional variant |
| Outcome | Linkage disequilibrium score as represented by r^2^ |

**Supplementary Table S2.** Search queries performed for the identification of studies.

| Resource | Search Strategy | Numbe of Results |
| --- | --- | --- |
| PubMed/Medline (National Library of Medicine, 1946-2022) | ((("ABO Blood-Group System"[MESH]) OR ("Blood Group System*"[ALL] AND (ABO[ALL] OR ABH[ALL] OR H[ALL] OR "H type 1 Antigen"[ALL])) OR ("Blood-Group System*"[ALL] AND (ABO[ALL] OR ABH[ALL] OR H[ALL] OR "H type 1 Antigen"[ALL])) OR ("Blood Factor*"[ALL] AND (ABO[ALL] OR ABH[ALL] OR H[ALL] OR "H type 1 Antigen"[ALL])) OR ("Blood Type A"[ALL]) OR ("Blood Type"[ALL] AND (B[ALL] OR O[ALL] OR AB[ALL])) OR "ABO Factor*" OR ("ABO Blood Group Antigen*"[ALL])) AND ("Polymorphism, Single Nucleotide"[MESH] OR "Genetic Linkage"[Mesh] OR "Single Nucleotide Polymorphism*"[ALL] OR "Linkage Disequilibrium"[Mesh] OR SNP*[ALL] OR "Tagging SNP*"[ALL] OR "Tag SNP*"[ALL] OR tSNP*[ALL] OR "Genetic Linkage"[ALL] OR "Genetic Linkage Analysis" [ALL] OR "Genetic Linkage Analyses" [ALL] OR "Linkage Disequilibrium*" [ALL] OR "LD score*" [ALL] OR "Linkage" [ALL])) NOT ((review[Publication Type]) OR ("letter"[Publication Type] OR "editorial"[Publication Type])) Filters: Humans | 421 |
| Embase (Elsevier, 1947-2022) | 'single nucleotide polymorphism'/exp OR 'genetic linkage'/exp OR 'gene linkage disequilibrium'/exp OR 'linkage disequilibrium' OR 'snp*' OR 'taging snp*' OR 'tag snp*' OR 'tsnp*' OR 'genetic linkage' OR 'genetic linkage analysis' OR 'genetic linkage analyses' OR 'ld score*'  AND  'blood group abo system'/exp OR 'blood group system'/exp OR 'abh blood group system*' OR 'h blood group system*' OR 'h type 1 antigen blood group system' OR 'blood group'/exp OR 'blood factor' OR 'abo blood factor*' OR 'abh blood factor*' OR 'h blood factor*' OR 'h type 1 antigen blood factor*' OR 'blood group a'/exp OR 'blood type a' OR 'blood group b'/exp OR 'blood type b' OR 'blood group o'/exp OR 'blood type o' OR 'abo factor*' OR 'abo blood group antigen'  AND  ([article]/lim OR [article in press]/lim) AND [humans]/lim AND [english]/lim | 823 |
| Scopus (Elsevier, 1788-2022) | ( TITLE-ABS-KEY ( "Single Nucleotide Polymorphism*" OR "SNP*" OR "Tag SNP*" OR "Tag-SNP*" OR "Tagging SNP*" OR "tSNP*" OR "Genetic linkage" OR "LD Score" OR "Linkage Disequilibrium" OR "Genetic Linkage Analysis" OR "Genetic Linkage Analyses" ) ) AND ( TITLE-ABS-KEY ( "ABO Blood-Group System*" OR "ABO Blood Group System*" OR "abh blood group*" OR "ABO factor*" OR "Blood Group H Type 1 Antigen" OR "h blood group*" OR "h blood group system" OR "blood type A" OR "blood type B" OR "blood type O" OR "blood type AB" ) ) AND ( LIMIT-TO ( DOCTYPE , "ar" ) ) AND ( LIMIT-TO ( EXACTKEYWORD , "Human" ) ) AND ( LIMIT-TO ( LANGUAGE , "English" ) ) | 293 |
| Ovid/Medline (Wolters Kluwer, 1946-2022) | exp ABO Blood-Group System/ or "ABO Blood-Group System*".ti,ab,kf. or "ABO Blood Group System*".ti,ab,kf. or "abh blood group*".ti,ab,kf. or "ABO factor*".ti,ab,kf. or "Blood ADJ Group ADJ H ADJ Type ADJ 1 ADJ Antigen".ti,ab,kf. or "h ADJ blood group*".ti,ab,kf. or "h ADJ blood group system".ti,ab,kf. or ("blood type ADJ A" or "blood type ADJ B" or "blood type ADJ O" or "blood type ADJ AB").ti,ab,kf.  AND  exp Polymorphism, Single Nucleotide/ or exp Genetic Linkage/ or exp Linkage Disequilibrium/ or SNP*.ti,ab,kf. or Single Nucleotide Polymorphism*.ti,ab,kf. or Genetic Linkage.ti,ab,kf. or Genetic Linkage Analysis.ti,ab,kf. or Genetic Linkage Analyses.ti,ab,kf. or Tag SNP*.ti,ab,kf. or tagging SNP*.ti,ab,kf. or tSNP*.ti,ab,kf. or Tag-SNP*.ti,ab,kf. or tag SNP*.ti,ab,kf. or LD Score.ti,ab,kf.  limit to (english language and humans) | 330 |
| CINHAL Plus with Full Text (Ebsco, 1937-2022) | (MH "ABO Blood-Group System") OR (MH "Blood Groups") OR (( "blood group system*" OR "blood-group system*" OR "blood factor" ) AND ( "ABO" OR "ABH" OR "H" OR "H type 1 antigen" )) OR (("blood type" AND ( "A" OR "B" OR "AB" OR "O" )) OR ("ABO factor*" OR "ABO Blood Group Antigen*")  AND  (MH "Polymorphism, Single Nucleotide") OR "genetic linkage" OR "Linkage Disequilibrium" OR SNP* OR "Tagging SNP*" OR "Tag SNP*" OR tSNP* OR "Genetic Linkage Analysis" OR "Genetic Linkage Analyses" OR "LD score*" | 32 |
| Google Scholar Advanced search | Find articles  with all of the words: SNP, abo  with the exact phrase: abo blood type  where my words occur: anywhere in the article | 726 |
| Dissertations and Theses Global | AB,TI("Single Nucleotide Polymorphism*" OR "SNP*" OR "Tag SNP*" OR "Tag-SNP*" OR "Tagging SNP*" OR "tSNP*" OR "Genetic linkage" OR "LD Score" OR "Linkage Disequilibrium" OR "Genetic Linkage Analysis" OR "Genetic Linkage Analyses") AND AB,TI("ABO Blood-Group System*" OR "ABO Blood Group System*" OR "abh blood group*" OR "ABO factor*" OR "Blood Group H Type 1 Antigen" OR "h blood group*" OR "h blood group system" OR "blood type A" OR "blood type B" OR "blood type O" OR "blood type AB") | 0 |

**Supplementary Table S3.** Initial screening form.

| **Question Text** | **Type** | **Answer Text** | **Inclusion Response*** |
| --- | --- | --- | --- |
| Is the associated article able to be found? | Radio | Yes, No | Yes |
| Is this reference in English? | Radio | Yes, No, Unclear | Yes, Unclear |
| Is the organism being studied human? | Radio | Yes, No, Unclear | Yes, Unclear |
| Is this a review article (describes previously published studies and does not describe original research results)? | Radio | Yes, No, Unclear | No, Unclear |
| Is this reference from a journal article describing original research and not from another source such as a textbook? | Radio | Yes, No, Unclear | Yes, Unclear |
| * This describes which answers allowed the reference to pass the initial screening and be included for subsequent evaluation. | | | |

**Supplementary Table S4.** Secondary screening and data extraction.

| **Question Text** | **Type** | **Answer Text** |
| --- | --- | --- |
| Are ABO blood (A, B, AB, O or A1, A2, A1A2, A1B, A2B, B, O)  groups being determined genetically using SNPs?***** | Radio | Yes, No |
| What SNP is being used to identify O alleles (rs ID is preferred, ex: rs8176719)? If no SNP being used answer as NA. | Text | Free Text |
| What SNP is being used to identify B alleles (rs ID is preferred, ex: rs8176746)? If no SNP being used answer as NA.****** | Text | Free Text |
| What SNP is being used to identify A alleles (rs ID is preferred)? If no SNP being used answer as NA. | Text | Free Text |
| What SNP is being used to identify A1 alleles (rs ID is preferred)? If no SNP being used answer as NA. | Text | Free Text |
| What SNP is being used to identify A2 alleles (rs ID is preferred, ex: rs56392308)? If no SNP being used answer as NA. | Text | Free Text |
| What number of SNPs are being used for ABO blood typing? | Radio | 0, 1, 2, 3, 4, 5, 5+ |
| Is a haplotype analysis performed, i.e. have they phased the data? | Radio | Yes, No, Unclear |
| Are gentetically derived blood groups compared to blood groups derived from serotyping? | Radio | Yes, No, Unclear |
| Are any LD scores reported? | Radio | Yes, No, Unclear |
| What LD or r2 scores are reported and between which SNPs? What is the reference population? | Text | Free Text |
| Does the study report a "statistically significant result" in relation to the ABO blood typing performed using SNPs? | Radio | Yes, No, Unclear |
| What is the total cohort size of the group that underwent ABO blood typing using SNPs (do not use commas)? | Text | Free Text |
| How is the population that was bloodtyped with SNPs described? | Text | Free Text |
| How wany individuals that can be described as African (AFR) were bloodtyped with SNPs? | Text | Free Text |
| How wany individuals that can be described as admixed American (AMR) were bloodtyped with SNPs? | Text | Free Text |
| How wany individuals that can be described as East Asian (EAS) were bloodtyped with SNPs? | Text | Free Text |
| How wany individuals that can be described as European (EUR) were bloodtyped with SNPs? | Text | Free Text |
| How wany individuals that can be described as South Asian (SAS) were bloodtyped with SNPs? | Text | Free Text |
| How wany individuals do not fit the description above and will be classified as "other" (often used for when studies refer to "Non-european/non-white other")? | Text | Free Text |
| What is the primary phenotype of interest? | Text | Free Text |
| How is the study type/design described? | Text | Free Text |
| What is the sequencing/genotyping platform? | Text | Free Text |
| * This describes another screening question where reviewers were prompted to include the reference and proceed with data extraction only if the response was “Yes”.  ** In colloquial terms we describe a B-defining SNP as the A blood type allele is considered the reference and the standard practice is to use a SNP with the alternative allele differentiating the B blood type from the A blood type. However, some studies will include both an A-defining SNP and a B-defining SNP. We sought to capture this nuance. In the absence of an A-defining SNP, we treating the “B-defining SNP” as an “A-B differentiating” SNP. | | |

**Supplementary Table S5.** Inferring ABO blood types using two functional variants.

| **Blood type allele** | **rs8176719** | **rs7853989** | **rs8176743** | **rs8176746** | **rs8176747** |
| --- | --- | --- | --- | --- | --- |
| A | C | G | C | G | C |
| O | - | * | * | * | * |
| B | C | C | T | T | G |
| * any allele | | | | | |

**Supplementary Table S6.** The extracted and derived data used for this study are available as a separate CSV document.

**Supplementary Table S7.** Journals in which included studies were published.

| **Journals** | **Number of Studies** |
| --- | --- |
| Journal of Thrombosis and Haemostasis | 14 |
| PLOS One | 12 |
| Human Molecular Genetics | 6 |
| Nature Genetics | 5 |
| Blood | 5 |
| International Journal of Cancer | 4 |
| Cancer Epidemiology; Biomarkers & Prevention | 4 |
| Transfusion | 3 |
| Cancer Medicine | 3 |
| BMC Genomics; Neurology; Annals of Human Genetics; Journal of Medical Genetics; Cancer Science; Blood Coagulation & Fibrinolysis; medRxiv _preprint; Legal Medicine; American Journal of Hematology; Nature Communications; Gut; Thrombosis Research; Arteriosclerosis; Thrombosis; and Vascular Biology | 2 |
| Malaria Journal; J Thromb Haemost; Annals of Neurology; World Journal of Gastroenterology; Blood Cells; Molecules and Diseases; International Journal of Cardiology; Psychiatry Research; BMC Medicine; Journal of Clinical Lipidology; TH Open; The Prostate; PLOS Genetics; Journal of Hepatology; Cancer Causes and Control; Infection; Genetics and Evolution; American Journal of Epidemiology; Oncology Reports; BMC Medical Genetics; The Journal of Clinical Endocrinology and Metabolism; Genes & Immunity; Communications Biology; International Journal of Immunogenetics; Journal of Gastroenterology and Hepatology; Thrombosis and Haemostasis; Carcinogenesis; Journal of Crohn's and Colitis; Journal of Clinical Laboratory Analysis; Epidemiology; Neurobiology of Aging; Journal of Bone and Joint Surgery; Transfusion Medicine and Hemotherapy; BMC Cancer; Frontiers in Medicine; Immunohematology; Diabetes; Pharmacogenetics and Genomics; Journal of Thrombosis and Thrombolysis; American Journal of Reproductive Immunology; New England Journal of Medicine; Journal of Clinical Pharmacy and Therapeutics; The Pharmacogenomics Journal; Journal of Infectious Diseases; Australian Journal of Forensic Sciences; npj Genomic Medicine; Journal of Forensic Sciences; Pancreas; Journal of Epidemiology; Cancer Research; Journal of Nutrition; Asian Pacific Journal of Cancer Prevention; Human Genome Variation; Central European Journal of Biology; PLOS Medicine; Aging Medicine | 1 |

**Supplementary Table S8.** Population breakdown across studies.

| **Population(s)** | **Count** | **Proportion** |
| --- | --- | --- |
| EUR | 59 | 0.433824 |
| EAS | 30 | 0.220588 |
| AFR | 7 | 0.051471 |
| AFR, EUR | 6 | 0.044118 |
| EUR, other | 4 | 0.029412 |
| AMR | 3 | 0.022059 |
| AFR, EUR, other | 2 | 0.014706 |
| AFR, AMR, EUR, other | 2 | 0.014706 |
| AFR, EAS, EUR, other | 2 | 0.007353 |
| SAS | 1 | 0.007353 |
| AFR, other | 1 | 0.007353 |
| unclear | 19 | 0.147509 |

**Supplementary Table S9.** SNP configurations for determining ABO bloodtype alleles.

| **O vs non-O SNPs** | **SNP configuration** | **Count** | **Percentage** |
| --- | --- | --- | --- |
|  | rs8176719 | 62 | 0.455882 |
|  | rs505922 | 28 | 0.205882 |
|  | rs687289 | 14 | 0.102941 |
|  | rs514659 | 4 | 0.029412 |
|  | rs8176719, rs505922 | 3 | 0.022059 |
|  | rs657152 | 2 | 0.014706 |
|  | rs8176645 | 2 | 0.014706 |
|  | rs8176719, rs687289 | 2 | 0.014706 |
|  | rs8176704 | 1 | 0.007353 |
|  | rs72238104 | 1 | 0.007353 |
|  | rs657152, rs612169 | 1 | 0.007353 |
|  | rs505922, rs612169 | 1 | 0.007353 |
|  | rs687289, rs505922 | 1 | 0.007353 |
|  | rs612169 | 1 | 0.007353 |
|  | rs529565, rs8176693 | 1 | 0.007353 |
|  | NA | 3 | 0.022059 |
|  | unclear | 9 | 0.066176 |
| **A vs B SNPs** | s8176746 | 54 | 0.397059 |
|  | rs8176749 | 13 | 0.095588 |
|  | rs8176746, rs8176747 | 13 | 0.095588 |
|  | rs8176747 | 10 | 0.073529 |
|  | rs8176743 | 5 | 0.036765 |
|  | rs8176672 | 2 | 0.014706 |
|  | rs7853989, rs8176720 | 1 | 0.007353 |
|  | rs8176747, rs8176720, rs8176741, rs8176746 | 1 | 0.007353 |
|  | rs8176746, rs8176749 | 1 | 0.007353 |
|  | rs8176746, rs8176672 | 1 | 0.007353 |
|  | rs8176747, rs7853989 | 1 | 0.007353 |
|  | rs8176746, rs8176743 | 1 | 0.007353 |
|  | rs8176722 | 1 | 0.007353 |
|  | rs8176720 | 1 | 0.007353 |
|  | rs7853989 | 1 | 0.007353 |
|  | rs8176743, rs8176746, rs8176749 | 1 | 0.007353 |
|  | NA | 19 | 0.139706 |
|  | unclear | 10 | 0.073529 |

**Supplementary Table S10.** Kruskal-Wallis Test for r^2^ across 1000 Genome Project continental populations.

| **statistic** | **parameter** | **p.value** | **tsnp** | **functional variant** |
| --- | --- | --- | --- | --- |
| 17.90598 | 4 | 0.001287** | rs505922 | rs8176719 |
| 18.99744 | 4 | 0.000787*** | rs514659 | rs8176719 |
| 18.05641 | 4 | 0.001203** | rs529565 | rs8176719 |
| 18.99744 | 4 | 0.000787*** | rs612169 | rs8176719 |
| 20.74243 | 4 | 0.000356*** | rs657152 | rs8176719 |
| 19.97264 | 4 | 0.000506*** | rs687289 | rs8176719 |
| 15.55253 | 4 | 0.003682** | rs8176645 | rs8176719 |
| 19.56215 | 4 | 0.000609*** | rs8176693 | rs8176719 |
| 8.916512 | 3 | 0.030422* | rs8176704 | rs8176719 |
| 16.76458 | 4 | 0.002147** | rs7853989 | rs8176746 |
| 18.9966 | 4 | 0.000787*** | rs8176672 | rs8176746 |
| 16.48706 | 4 | 0.002431** | rs8176720 | rs8176746 |
| 16.79641 | 4 | 0.002117** | rs8176722 | rs8176746 |
| 4.131124 | 4 | 0.388551 | rs8176741 | rs8176746 |
| 4.131124 | 4 | 0.388551 | rs8176743 | rs8176746 |
| 4.2 | 4 | 0.379615 | rs8176749 | rs8176746 |
| *p-value <0.05  **p-value <0.01  ***p-value <0.001 | | | | |

**Supplementary Table S11.** Dunn’s Test of multiple comparisons using rank sums across 1000 Genome Project continental populations.

| **pairwise_pop_comparisons** | **mean.rank.diff** | **pval** | **tsnp** | **functional variant** |
| --- | --- | --- | --- | --- |
| AMR-AFR | 9.5 | 0.475177 | rs505922 | rs8176719 |
| EAS-AFR | 16.6 | 0.002101** | rs505922 | rs8176719 |
| EUR-AFR | 10 | 0.255568 | rs505922 | rs8176719 |
| SAS-AFR | 15.2 | 0.006889** | rs505922 | rs8176719 |
| EAS-AMR | 7.1 | 1 | rs505922 | rs8176719 |
| EUR-AMR | 0.5 | 1 | rs505922 | rs8176719 |
| SAS-AMR | 5.7 | 1 | rs505922 | rs8176719 |
| EUR-EAS | -6.6 | 1 | rs505922 | rs8176719 |
| SAS-EAS | -1.4 | 1 | rs505922 | rs8176719 |
| SAS-EUR | 5.2 | 1 | rs505922 | rs8176719 |
| AMR-AFR | 9.25 | 0.536679 | rs514659 | rs8176719 |
| EAS-AFR | 17.6 | 0.00085*** | rs514659 | rs8176719 |
| EUR-AFR | 9.4 | 0.35825 | rs514659 | rs8176719 |
| SAS-AFR | 15 | 0.008101** | rs514659 | rs8176719 |
| EAS-AMR | 8.35 | 1 | rs514659 | rs8176719 |
| EUR-AMR | 0.15 | 1 | rs514659 | rs8176719 |
| SAS-AMR | 5.75 | 1 | rs514659 | rs8176719 |
| EUR-EAS | -8.2 | 0.900486 | rs514659 | rs8176719 |
| SAS-EAS | -2.6 | 1 | rs514659 | rs8176719 |
| SAS-EUR | 5.6 | 1 | rs514659 | rs8176719 |
| AMR-AFR | 9.5 | 0.475177 | rs529565 | rs8176719 |
| EAS-AFR | 16.4 | 0.002503** | rs529565 | rs8176719 |
| EUR-AFR | 9.8 | 0.286534 | rs529565 | rs8176719 |
| SAS-AFR | 15.6 | 0.004953** | rs529565 | rs8176719 |
| EAS-AMR | 6.9 | 1 | rs529565 | rs8176719 |
| EUR-AMR | 0.3 | 1 | rs529565 | rs8176719 |
| SAS-AMR | 6.1 | 1 | rs529565 | rs8176719 |
| EUR-EAS | -6.6 | 1 | rs529565 | rs8176719 |
| SAS-EAS | -0.8 | 1 | rs529565 | rs8176719 |
| SAS-EUR | 5.8 | 1 | rs529565 | rs8176719 |
| AMR-AFR | 9.25 | 0.536679 | rs612169 | rs8176719 |
| EAS-AFR | 17.6 | 0.00085*** | rs612169 | rs8176719 |
| EUR-AFR | 9.4 | 0.35825 | rs612169 | rs8176719 |
| SAS-AFR | 15 | 0.008101** | rs612169 | rs8176719 |
| EAS-AMR | 8.35 | 1 | rs612169 | rs8176719 |
| EUR-AMR | 0.15 | 1 | rs612169 | rs8176719 |
| SAS-AMR | 5.75 | 1 | rs612169 | rs8176719 |
| EUR-EAS | -8.2 | 0.900486 | rs612169 | rs8176719 |
| SAS-EAS | -2.6 | 1 | rs612169 | rs8176719 |
| SAS-EUR | 5.6 | 1 | rs612169 | rs8176719 |
| AMR-AFR | 10.625 | 0.257669 | rs657152 | rs8176719 |
| EAS-AFR | 7.8 | 0.797461 | rs657152 | rs8176719 |
| EUR-AFR | 14.3 | 0.013168* | rs657152 | rs8176719 |
| SAS-AFR | 18.8 | 0.000241*** | rs657152 | rs8176719 |
| EAS-AMR | -2.825 | 1 | rs657152 | rs8176719 |
| EUR-AMR | 3.675 | 1 | rs657152 | rs8176719 |
| SAS-AMR | 8.175 | 1 | rs657152 | rs8176719 |
| EUR-EAS | 6.5 | 1 | rs657152 | rs8176719 |
| SAS-EAS | 11 | 0.221551 | rs657152 | rs8176719 |
| SAS-EUR | 4.5 | 1 | rs657152 | rs8176719 |
| AMR-AFR | 9 | 0.60425 | rs687289 | rs8176719 |
| EAS-AFR | 17.9 | 0.00064*** | rs687289 | rs8176719 |
| EUR-AFR | 8.8 | 0.493829 | rs687289 | rs8176719 |
| SAS-AFR | 15.5 | 0.005371** | rs687289 | rs8176719 |
| EAS-AMR | 8.9 | 0.827545 | rs687289 | rs8176719 |
| EUR-AMR | -0.2 | 1 | rs687289 | rs8176719 |
| SAS-AMR | 6.5 | 1 | rs687289 | rs8176719 |
| EUR-EAS | -9.1 | 0.599019 | rs687289 | rs8176719 |
| SAS-EAS | -2.4 | 1 | rs687289 | rs8176719 |
| SAS-EUR | 6.7 | 1 | rs687289 | rs8176719 |
| AMR-AFR | 11.125 | 0.200933 | rs8176645 | rs8176719 |
| EAS-AFR | 12.5 | 0.051758 | rs8176645 | rs8176719 |
| EUR-AFR | 15.3 | 0.006213** | rs8176645 | rs8176719 |
| SAS-AFR | 12.7 | 0.045026* | rs8176645 | rs8176719 |
| EAS-AMR | 1.375 | 1 | rs8176645 | rs8176719 |
| EUR-AMR | 4.175 | 1 | rs8176645 | rs8176719 |
| SAS-AMR | 1.575 | 1 | rs8176645 | rs8176719 |
| EUR-EAS | 2.8 | 1 | rs8176645 | rs8176719 |
| SAS-EAS | 0.2 | 1 | rs8176645 | rs8176719 |
| SAS-EUR | -2.6 | 1 | rs8176645 | rs8176719 |
| AMR-AFR | -6.42857 | 1 | rs8176693 | rs8176719 |
| EAS-AFR | 6.371429 | 1 | rs8176693 | rs8176719 |
| EUR-AFR | -9.22857 | 0.393384 | rs8176693 | rs8176719 |
| SAS-AFR | 8.371429 | 0.615894 | rs8176693 | rs8176719 |
| EAS-AMR | 12.8 | 0.126048 | rs8176693 | rs8176719 |
| EUR-AMR | -2.8 | 1 | rs8176693 | rs8176719 |
| SAS-AMR | 14.8 | 0.039198* | rs8176693 | rs8176719 |
| EUR-EAS | -15.6 | 0.012602* | rs8176693 | rs8176719 |
| SAS-EAS | 2 | 1 | rs8176693 | rs8176719 |
| SAS-EUR | 17.6 | 0.002744** | rs8176693 | rs8176719 |
| AMR-AFR | -0.92857 | 1 | rs8176704 | rs8176719 |
| EUR-AFR | 2.971429 | 1 | rs8176704 | rs8176719 |
| SAS-AFR | -8.22857 | 0.141135 | rs8176704 | rs8176719 |
| EUR-AMR | 3.9 | 1 | rs8176704 | rs8176719 |
| SAS-AMR | -7.3 | 0.476763 | rs8176704 | rs8176719 |
| SAS-EUR | -11.2 | 0.025902* | rs8176704 | rs8176719 |
| AMR-AFR | -11.6071 | 0.134193 | rs7853989 | rs8176746 |
| EAS-AFR | 3.942857 | 1 | rs7853989 | rs8176746 |
| EUR-AFR | -11.0571 | 0.11696 | rs7853989 | rs8176746 |
| SAS-AFR | -1.05714 | 1 | rs7853989 | rs8176746 |
| EAS-AMR | 15.55 | 0.01969* | rs7853989 | rs8176746 |
| EUR-AMR | 0.55 | 1 | rs7853989 | rs8176746 |
| SAS-AMR | 10.55 | 0.35752 | rs7853989 | rs8176746 |
| EUR-EAS | -15 | 0.015428* | rs7853989 | rs8176746 |
| SAS-EAS | -5 | 1 | rs7853989 | rs8176746 |
| SAS-EUR | 10 | 0.347722 | rs7853989 | rs8176746 |
| AMR-AFR | 14.125 | 0.024197* | rs8176672 | rs8176746 |
| EAS-AFR | 11 | 0.114544 | rs8176672 | rs8176746 |
| EUR-AFR | 17.5 | 0.000575*** | rs8176672 | rs8176746 |
| SAS-AFR | 9.6 | 0.273344 | rs8176672 | rs8176746 |
| EAS-AMR | -3.125 | 1 | rs8176672 | rs8176746 |
| EUR-AMR | 3.375 | 1 | rs8176672 | rs8176746 |
| SAS-AMR | -4.525 | 1 | rs8176672 | rs8176746 |
| EUR-EAS | 6.5 | 1 | rs8176672 | rs8176746 |
| SAS-EAS | -1.4 | 1 | rs8176672 | rs8176746 |
| SAS-EUR | -7.9 | 0.927179 | rs8176672 | rs8176746 |
| AMR-AFR | -10.3929 | 0.301661 | rs8176720 | rs8176746 |
| EAS-AFR | 4.057143 | 1 | rs8176720 | rs8176746 |
| EUR-AFR | -2.94286 | 1 | rs8176720 | rs8176746 |
| SAS-AFR | 9.057143 | 0.4314 | rs8176720 | rs8176746 |
| EAS-AMR | 14.45 | 0.048576* | rs8176720 | rs8176746 |
| EUR-AMR | 7.45 | 1 | rs8176720 | rs8176746 |
| SAS-AMR | 19.45 | 0.001501** | rs8176720 | rs8176746 |
| EUR-EAS | -7 | 1 | rs8176720 | rs8176746 |
| SAS-EAS | 5 | 1 | rs8176720 | rs8176746 |
| SAS-EUR | 12 | 0.131127 | rs8176720 | rs8176746 |
| AMR-AFR | -1.28571 | 1 | rs8176722 | rs8176746 |
| EAS-AFR | 14.11429 | 0.015648* | rs8176722 | rs8176746 |
| EUR-AFR | -0.48571 | 1 | rs8176722 | rs8176746 |
| SAS-AFR | 9.314286 | 0.368958 | rs8176722 | rs8176746 |
| EAS-AMR | 15.4 | 0.02597* | rs8176722 | rs8176746 |
| EUR-AMR | 0.8 | 1 | rs8176722 | rs8176746 |
| SAS-AMR | 10.6 | 0.381668 | rs8176722 | rs8176746 |
| EUR-EAS | -14.6 | 0.024573* | rs8176722 | rs8176746 |
| SAS-EAS | -4.8 | 1 | rs8176722 | rs8176746 |
| SAS-EUR | 9.8 | 0.420664 | rs8176722 | rs8176746 |
| *p-value <0.05  **p-value <0.01  ***p-value <0.001 | | | | |

**Supplementary Table S12.** Kruskal-Wallis and Dunn’s Test for multiple comparisons using ranks sums across inferred ancestral groups for r^2^ values for studies that only used tSNPs.

| **O vs non-O**  **SNPs** | **Kruskal-Wallis** | chi-squared = 27.455, df = 3, p-value = 4.727e-06 | | |
| --- | --- | --- | --- | --- |
|  | **Dunn’s Test** | **Pairwise Inferred Continental Ancestry Comparisons** | **Mean Rank Difference** | **P-Value** |
|  |  | AMR-AFR | 2.50000 | 1.0000 |
|  |  | EAS-AFR | 41.60000 | 1.1e-05 |
|  |  | EUR-AFR | 22.41892 | 0.0230 |
|  |  | EAS-AMR | 39.10000 | 0.0682 |
|  |  | EUR-AMR | 19.91892 | 1.0000 |
|  |  | EUR-EAS | -19.18108 | 0.0016 |
| **A vs B**  **SNPs** | **Kruskal-Wallis** | chi-squared = 8.6223, df = 2, p-value = 0.01342 | | |
|  | **Dunn’s Test** | **Pairwise Inferred Continental Ancestry Comparisons** | **Mean Rank Difference** | **P-Value** |
|  |  | AMR-AFR | -8.0000000 | 0.0272 |
|  |  | EAS-AFR | -0.7916667 | 1.0000 |
|  |  | EUR-AFR | 7.2083333 | 0.0149 |

**Supplementary Table S13.** Fisher’s exact test adjusted for pairwise comparisons for proportion of individuals that have inappropriate tSNPs.

|  | **Inferred Continental Ancestry Comparisons** | **N** | **P-value** | **P-value adjusted** |
| --- | --- | --- | --- | --- |
| **O vs non-O**  **SNPs** | AFR-AMR | 8311 | 1 | 1 |
|  | AFR-EAS | 139527 | 0 | 0 |
|  | AFR-EUR | 659533 | 0 | 0 |
|  | AMR-EAS | 135370 | 0 | 0 |
|  | AMR-EUR | 655376 | 0 | 0 |
|  | EAS-EUR | 786592 | 0 | 0 |
| **A vs B**  **SNPs** | AFR-EAS | 6372 | 1.99e-144 | 3.98e-144 |
|  | AFR-EUR | 74072 | 1.35e-168 | 4.05e-168 |
|  | EAS-EUR | 68142 | 3.79e- 50 | 3.79e- 50 |

**Supplementary Table S14.** Fisher’s exact test adjusted for pairwise comparisons for proportion of studies that have inappropriate tSNPs.

|  | **Inferred Continental Ancestry Comparisons** | **N** | **P-value** | **P-value adjusted** |
| --- | --- | --- | --- | --- |
| **O vs non-O**  **SNPs** | AFR-AMR | 5 | 1 | 1 |
|  | AFR-EAS | 14 | 0.000999 | 0.005 |
|  | AFR-EUR | 41 | 1 | 1 |
|  | AMR-EAS | 11 | 0.0909 | 0.364 |
|  | AMR-EUR | 38 | 1 | 1 |
|  | EAS-EUR | 47 | 0.000000193 | 0.00000116 |
| **A vs B**  **SNPs** | AFR-EAS | 5 | 0.4 | 0.825 |
|  | AFR-EUR | 15 | 1 | 1 |
|  | EAS-EUR | 14 | 0.275 | 0.825 |

**Supplementary Table S15.** Descriptive statistics for r^2^ values between tSNPs and functional variant values across 1000 Genomes Project-defined superpopulations are available as a separate CSV document.

**SUPPLEMENTARY FIGURES**

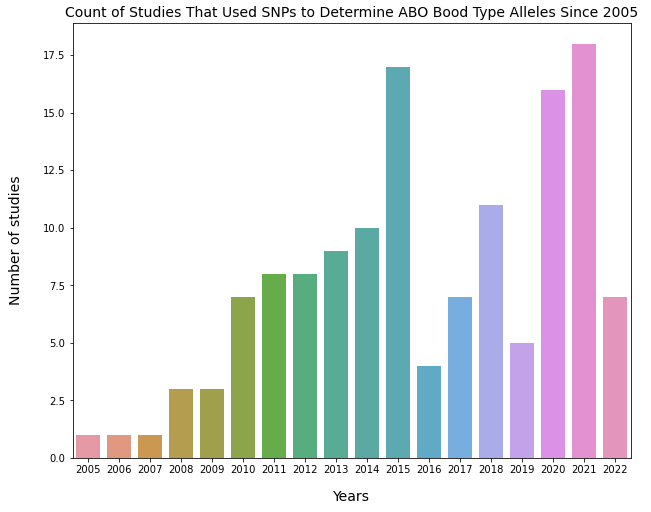

**Supplementary Figure S1. Count of studies that used SNPs to determine ABO alleles since 2005.** The numbers of studies that were published each year between 2005 and 2022 were counted. The count of studies each year is the following (year=count of studies): 2005=1, 2006=1, 2007=1, 2008=3, 2009=3, 2010=7, 2011=8, 2012=8, 2013=9, 2014=10, 2015=17, 2016=6, 2017=7, 2018=11, 2019=5, 2020=16, 2021=18, 2022=7.

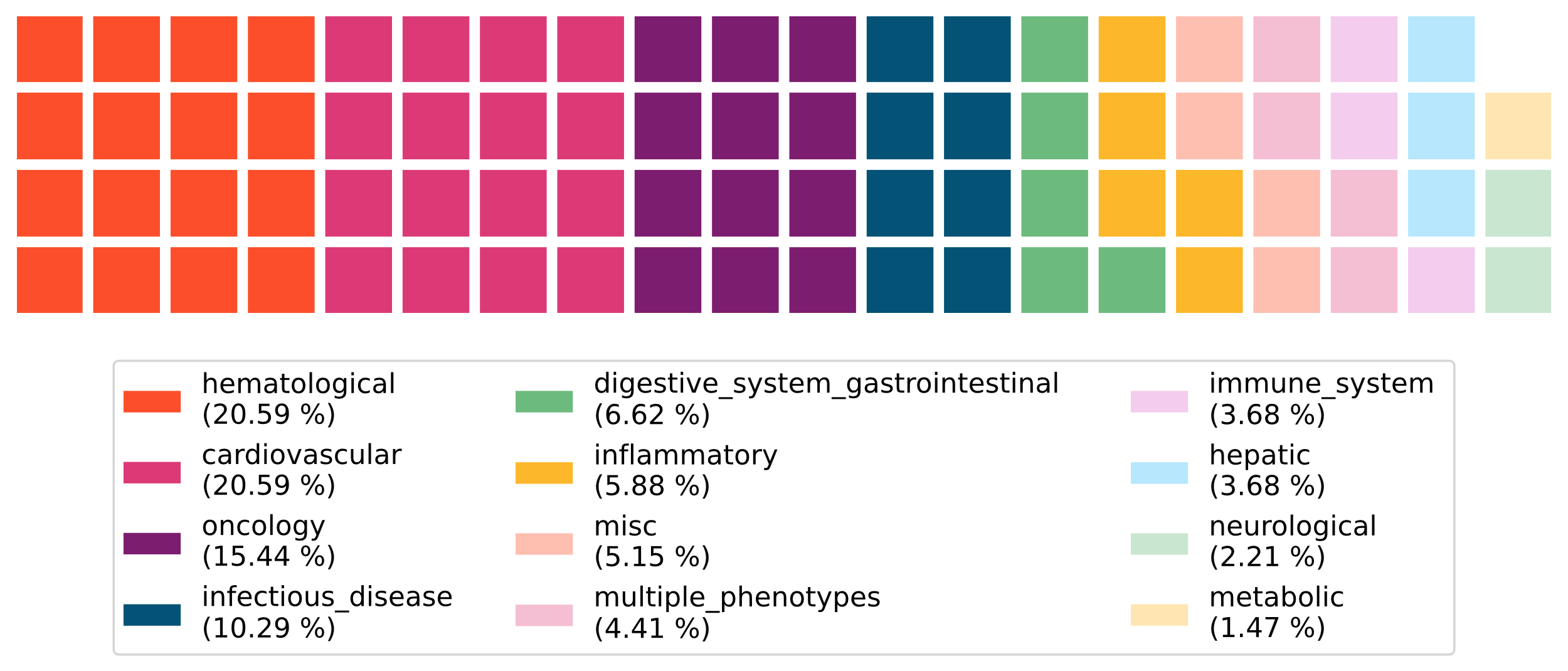

**Supplementary Figure S2. Primary phenotypes of the included studies.** The primary phenotype being investigated by each study was categorized into one of twelve categories and the proportion was then calculated. The counts are the following (phenotype=count of studies): hematological=28, cardiovascular=27, oncology=21, infectious disease=14, digestive system/gastrointestinal=9, inflammatory=8, miscilleanous=7, multiple phenotypes=6, immune system=5, hepatic=5, neurological=3, metabolic=2.

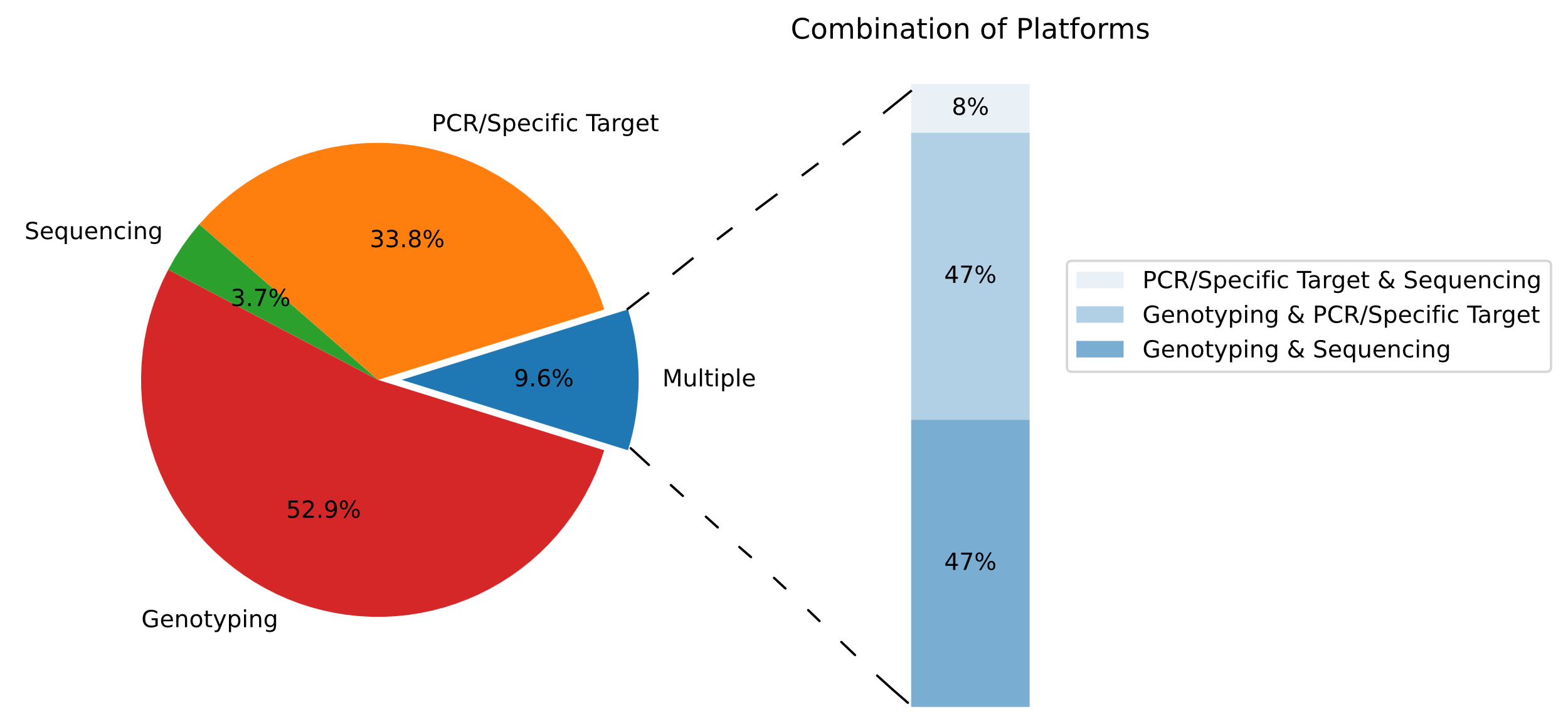

**Supplementary Figure S3. Genotyping/sequencing platforms used by included studies.** The sequencing/genotyping platforms used by studies were categorized into four groups: 1) sequencing, 2) genotyping, 3) PCR/specific target, and 4) multiple platforms. Genotyping describes platforms that do not sequence the whole genome but instead provides information on hundreds of thousands of DNA variants and includes data derived from microarray approaches. PCR/specific target describes platforms that genotype individuals for a small number of DNA variants. The multiple category represents studies that utilized more than on genotyping/sequencing platform. The counts per each category are the following (category=count of studies): genotyping=72, PCR/specific target=46, sequencing=5, and multiple platforms=13.

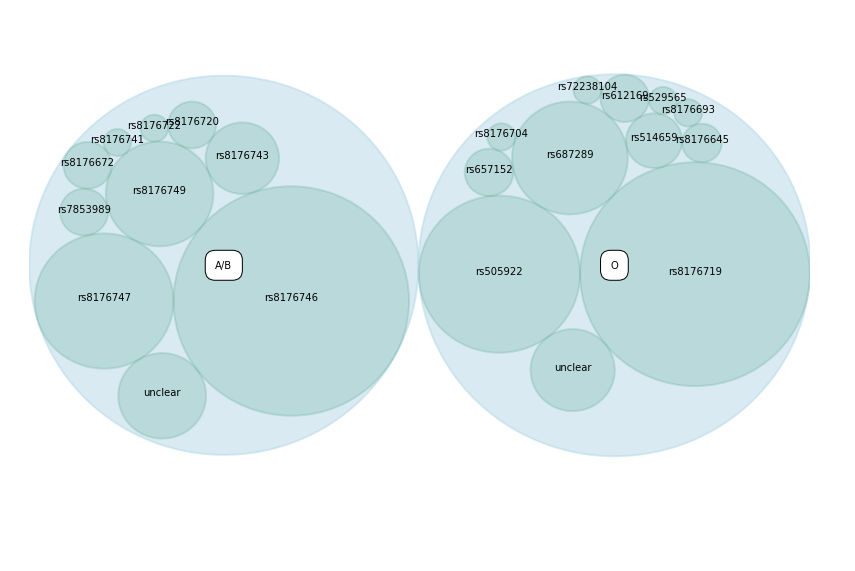

**Supplementary Figure S4. Hierarchical clustering plot for SNPs used to determine ABO alleles.** Plot represents the frequency of use of SNPs across studies. The functional variant that differentiate between O vs non-O alleles is rs8176719. The functional variants that defferentiate A vs B alleles are rs7853989, rs8176743, rs8176746, and rs8176747 .
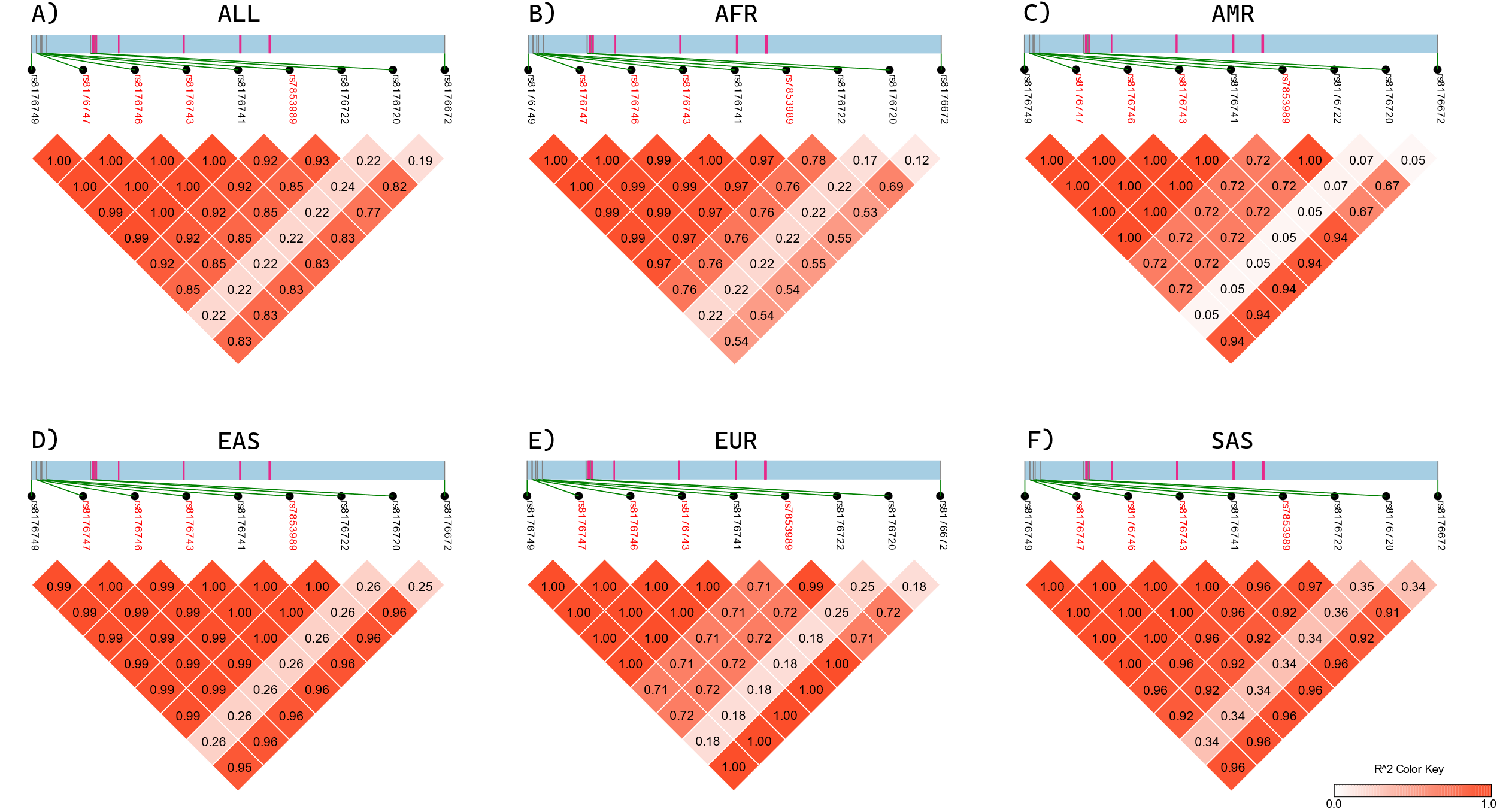

**Supplementary Figure S5.** **LD heatmap for SNPs that differentiate A vs B alleles across 1000 Genomes Project-defined superpopulations.** Figure represents a 10.97 kb region of the ABO gene starting at 133.256 Mb to 133.267 Mb. This region encompasses the locations of the A vs B functional variant and tSNPs. The blue block represents the noncoding regions and the pink lines represent exons. The functional variants are highlighted in red. Linkage dissociation blocks representing the r^2^ values of the A vs B functional variants and tSNPs are shown beneath the section of the ABO gene. **A)** Represents r^2^ values across all continental populations. **B) - F)** Represents r^2^ values across specific continental populations. AFR: Africa, AMR: America, EAS: East Asia, EUR: Europe, SAS: South Asia.
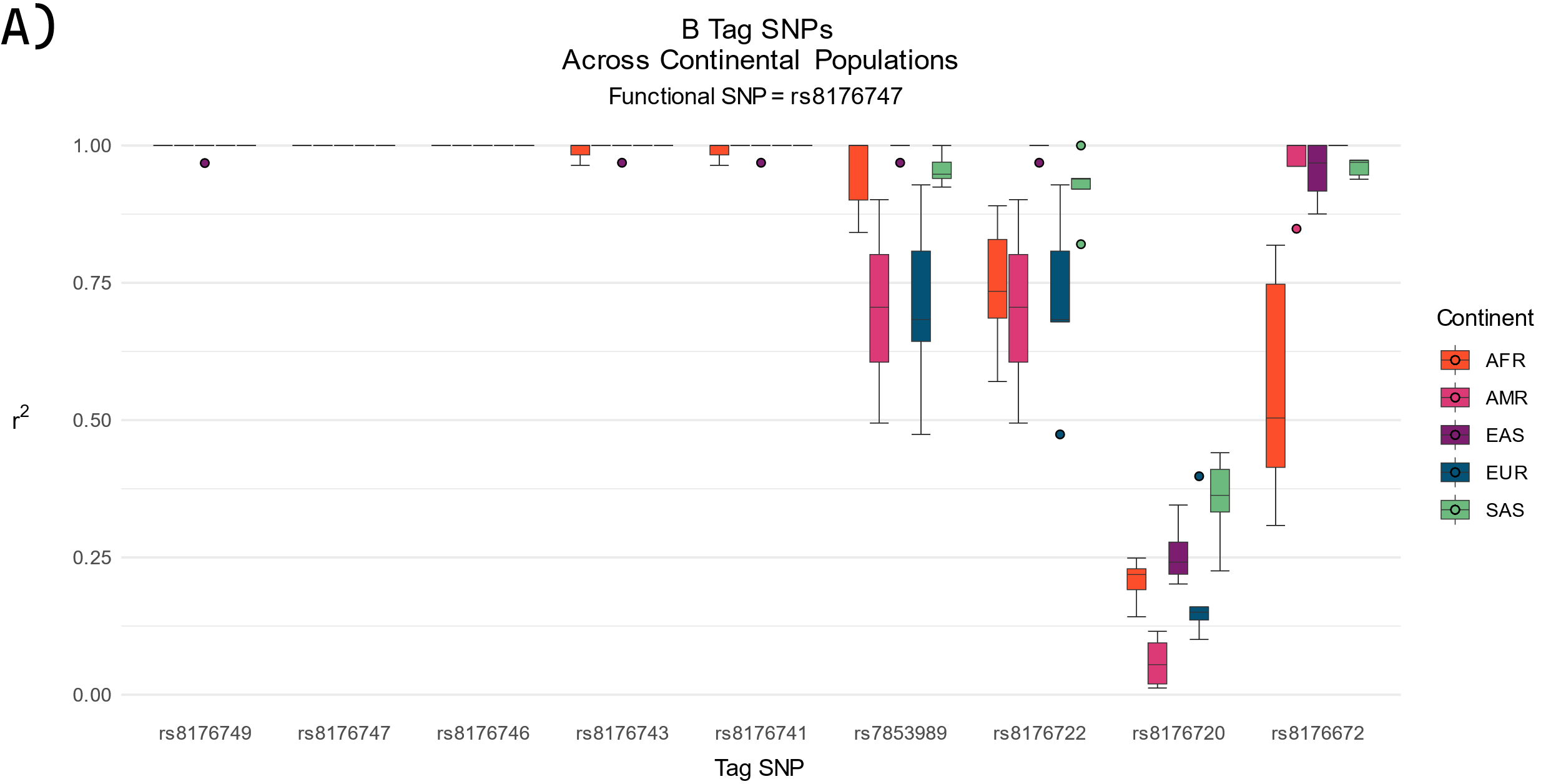

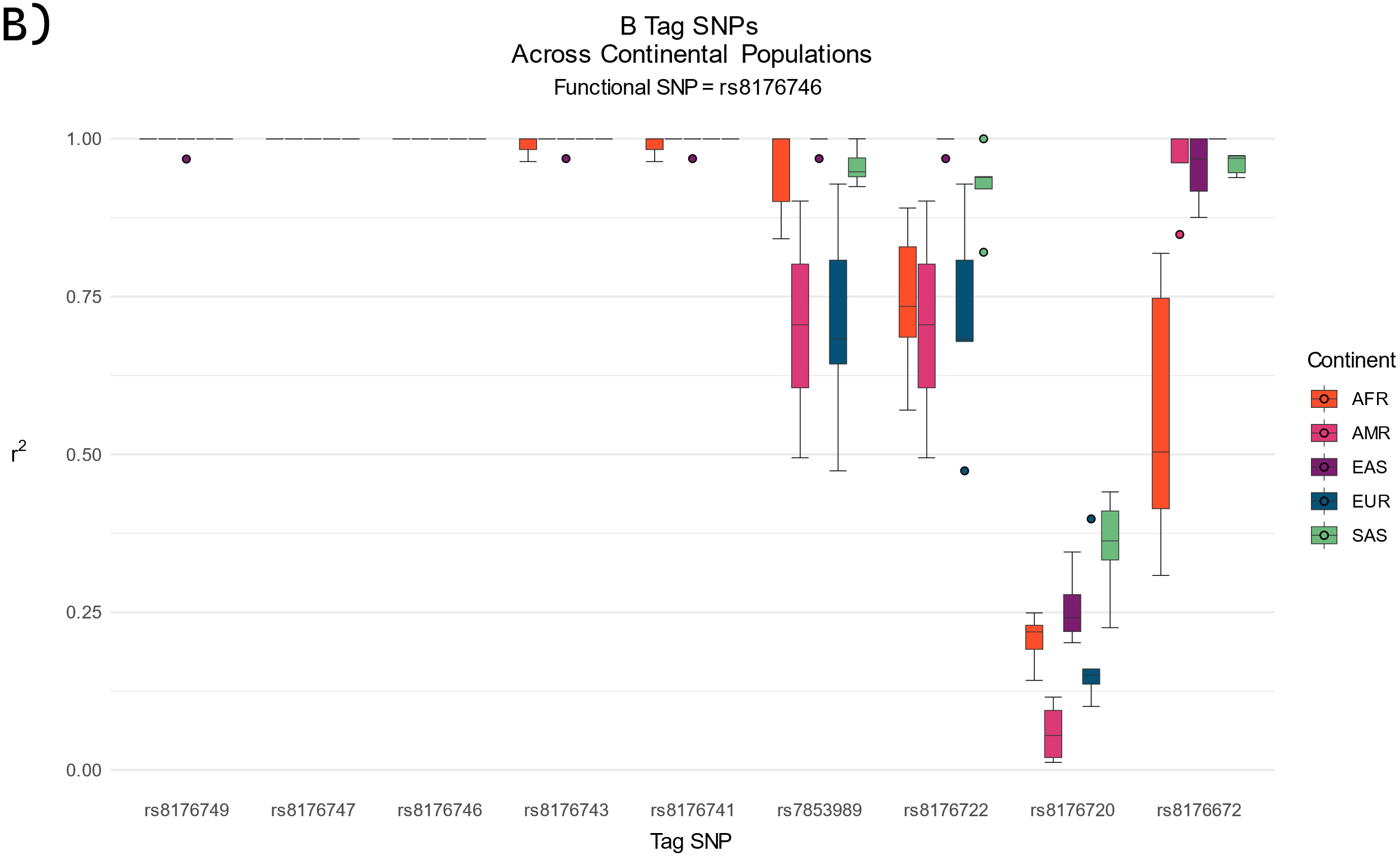

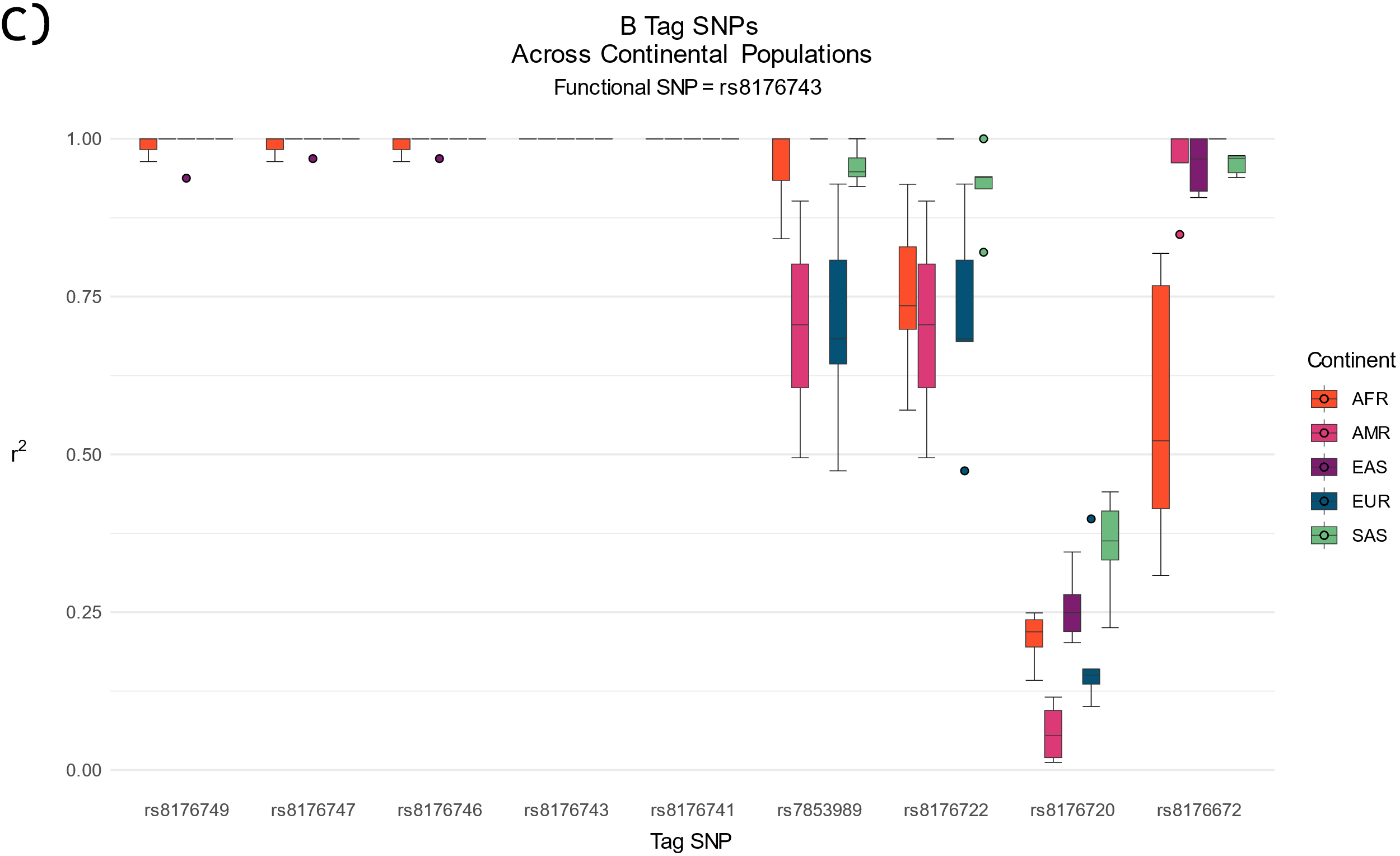

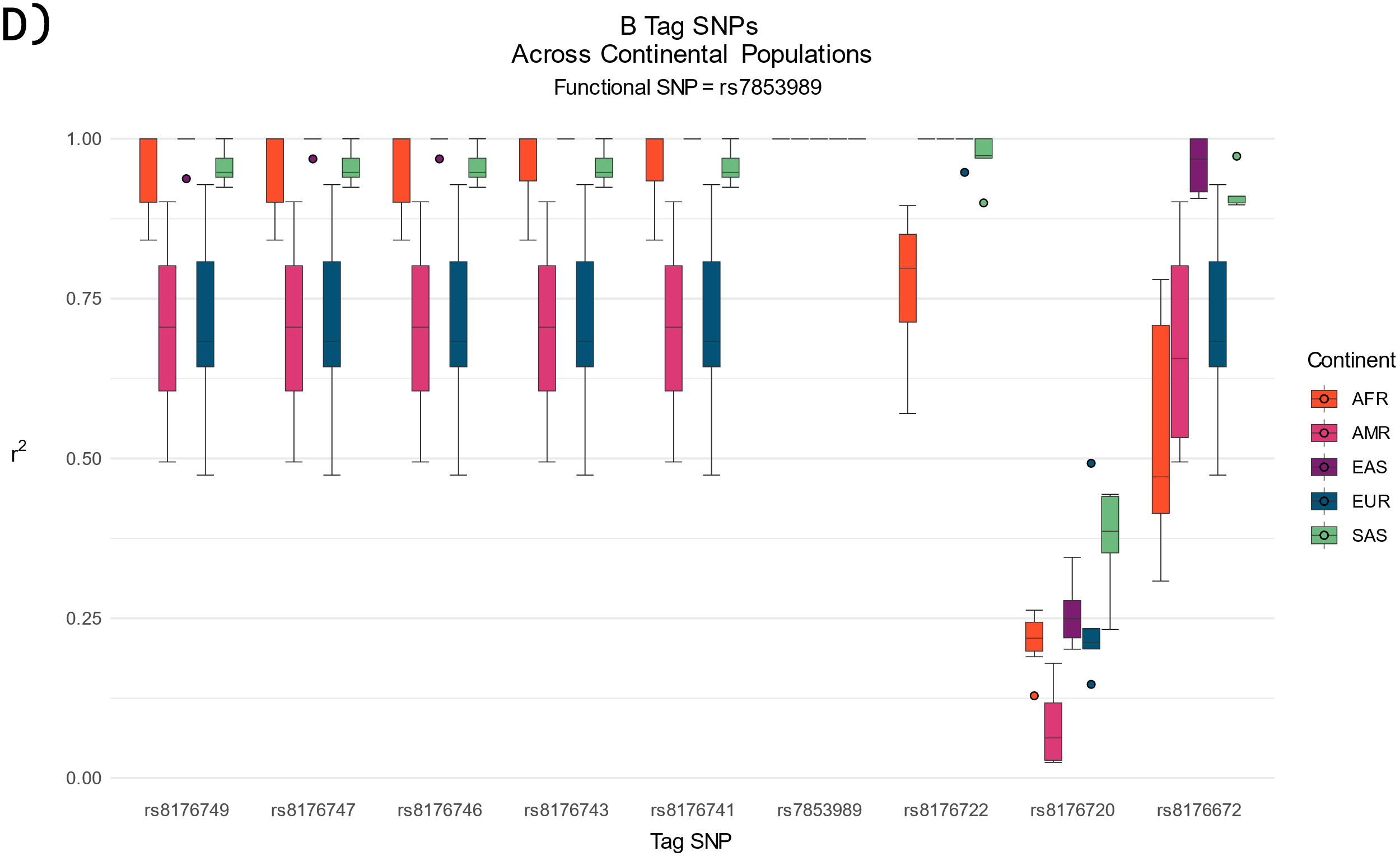

**Supplementary Figure S6**. **Distributions of r^2^ values for each 1000 Genomes Project-defined superpopulation per A vs B tSNP and functional variant**. **A)** The functional variant used for the calculation of r^2^ values is rs8176747. **B)** The functional variant used for the calculation of r^2^ values is rs8176746. **C)** The functional variant used for the calculation of r^2^ values is rs8176743. **D)** The functional variant used for the calculation of r^2^ values is rs7853989.

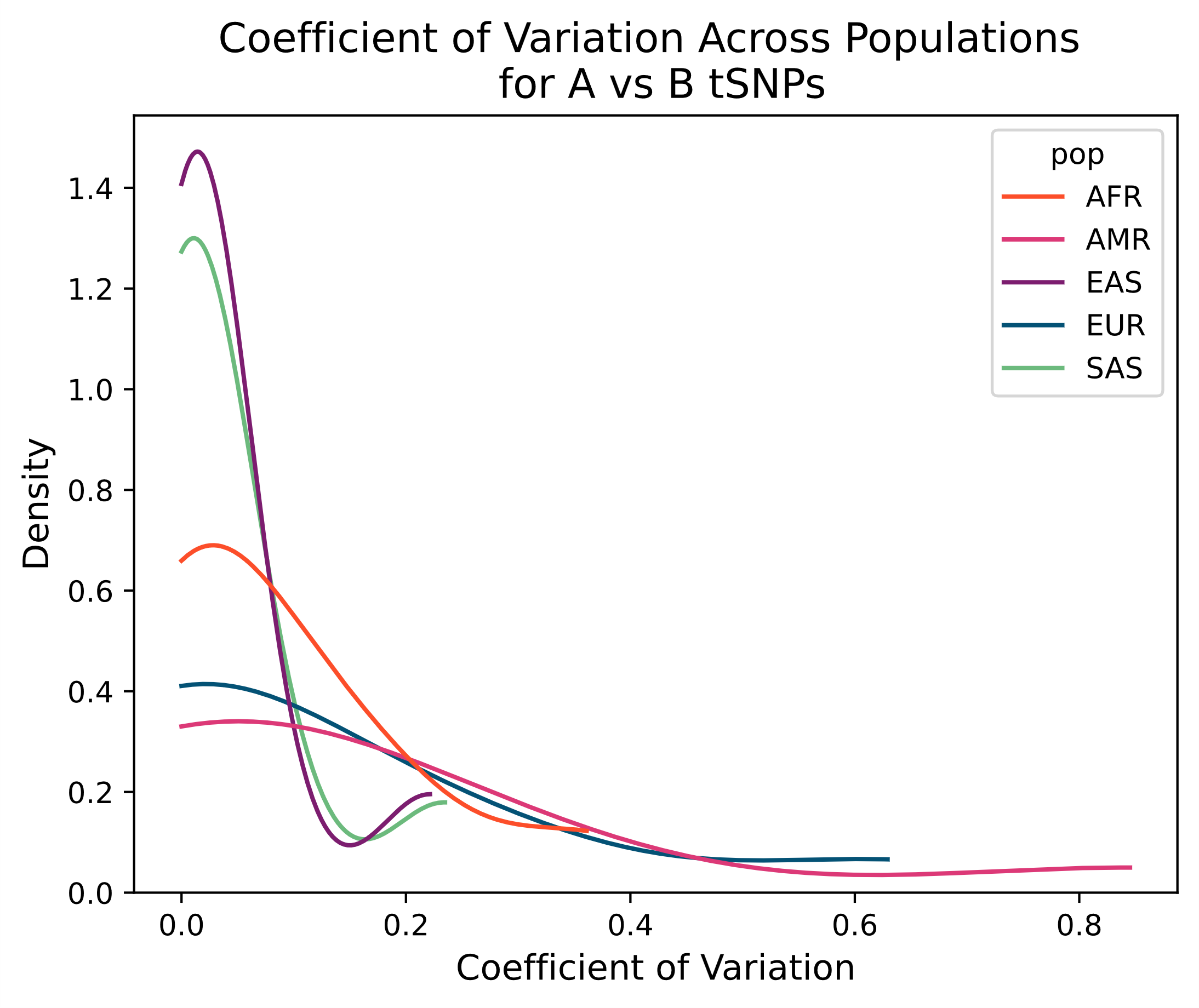

**Supplementary Figure S7.** **Plot of coefficient of determination.** The coefficient of determination for r^2^ between the A vs B functional variant and A vs B tSNPs was calculated for each subpopulation population and then plotted as a kernel density estimate plot across superpopulations (AFR, AMR, EAS, EUR, SAS).

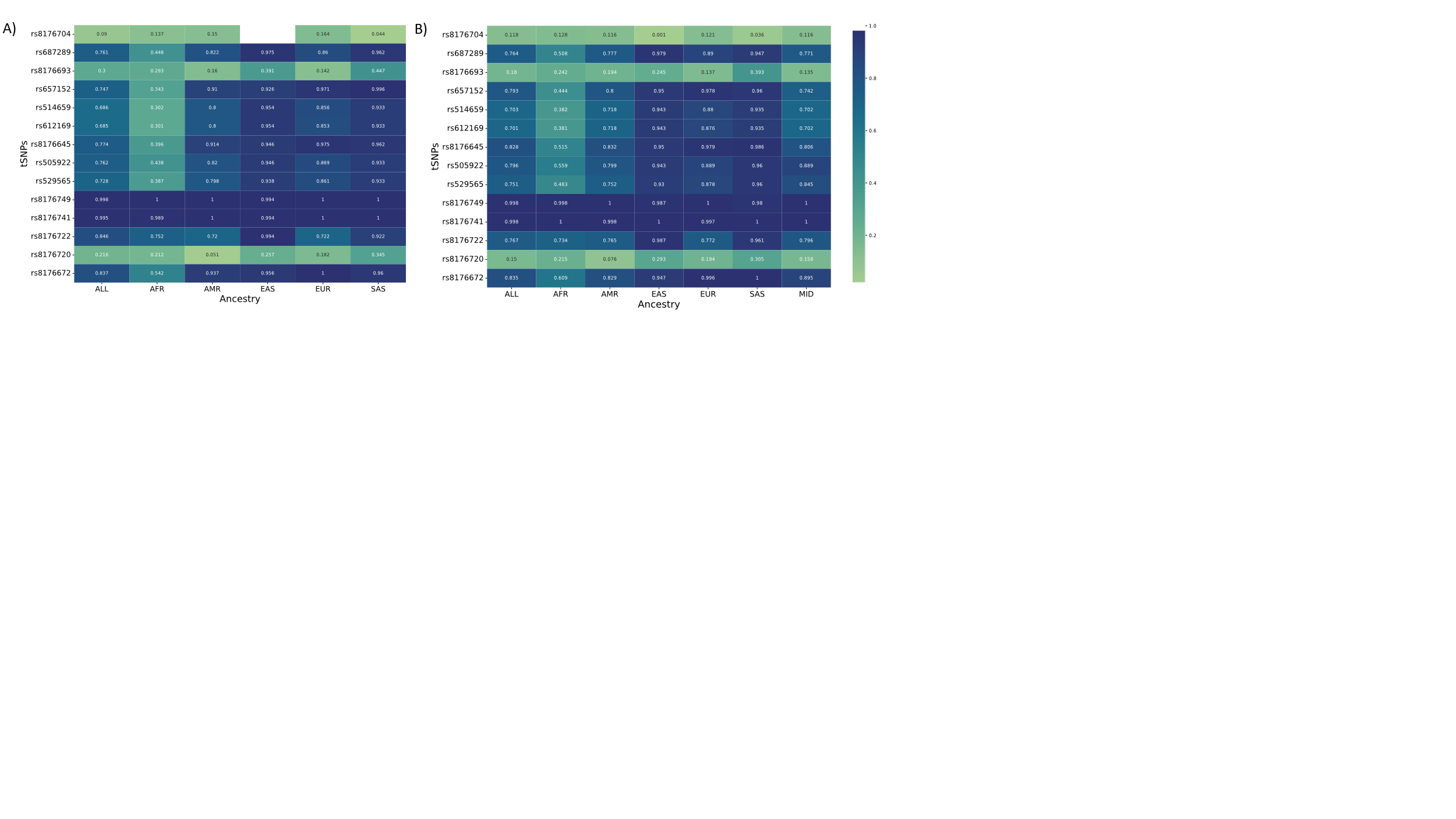

**Supplementary Figure S8.** **Linkage Disequilibirum (r^2^) between functional variants and tSNPs in 1000 Genomes Project cohort (A) and the *All of Us* cohort (B).** LD, reprented by r^2^ values, between the O vs non-O differentiating functional varaint (rs8176719) and O vs non-O differentiating tSNPs, and the A vs B differentiating functional variant (rs8176746) and A vs B tSNPs are displayed as a heatmap. r^2^ is calculated between rs8176719 and the following tSNPs: rs8176704, rs687289, rs8176693, rs657152, rs514659, rs612169, rs8176645, rs505922, and rs529565. r^2^ is calculated between rs8176719 and the following tSNPs: rs8176749, rs8176741, rs8176722, rs8176720, and rs8176672. LD was calculated in two separate cohorts: **A)** the 1000 Genomes Project 30x cohort (n=3,202) and **B)** the *All of Us* cohort (n=10,771).

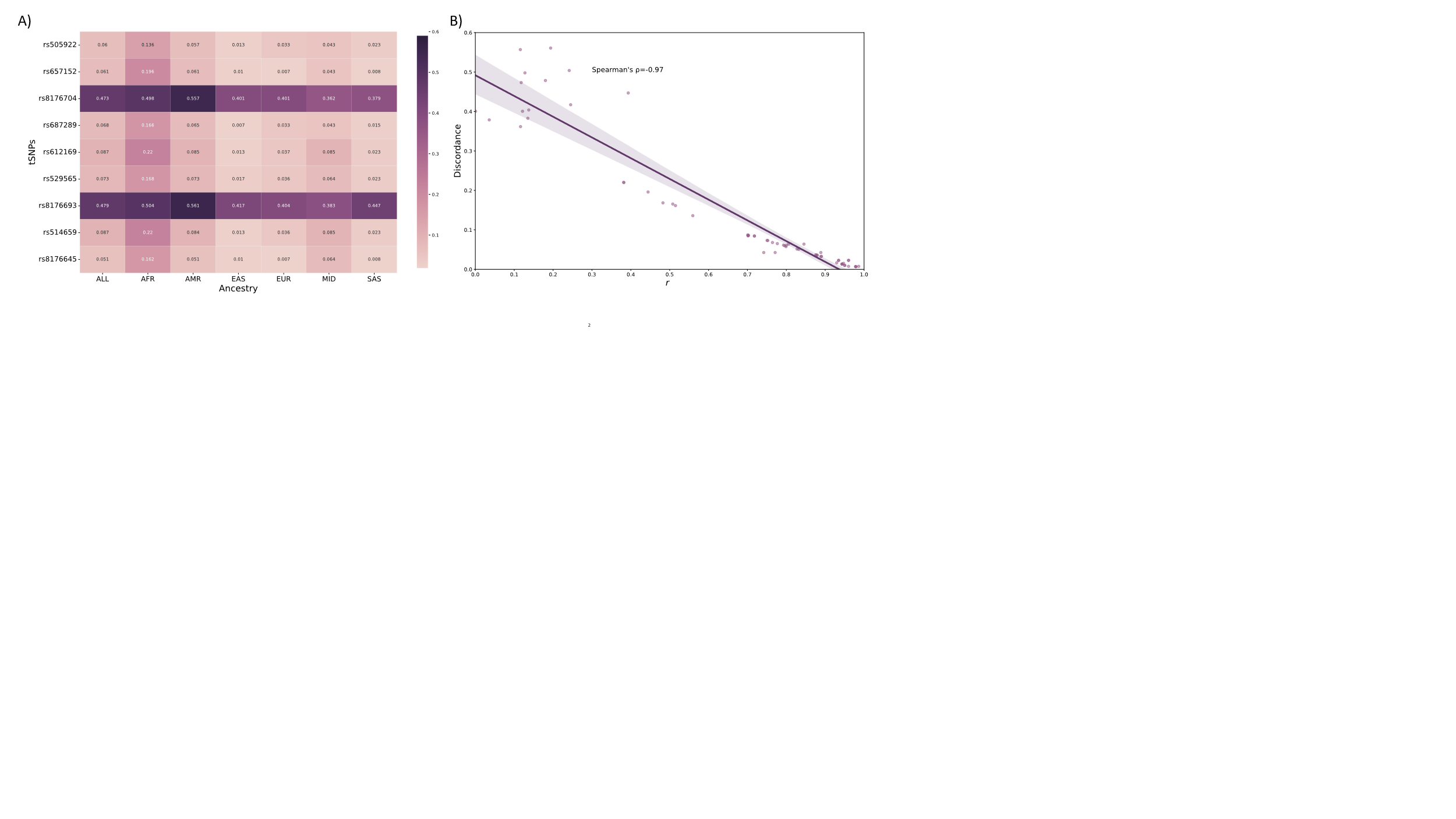

**Supplementary Figure S9.** **Discordance between O vs non-O blood types from tSNPs versus functional variants. A)** Discordance between ABO O and non-O blood types derived from the O vs non-O functional tSNPs and those derived from functional variants are displayed as a heatmap. Discordance (proportion of non-matches in a cohort of 10,771) was calculated across all ancestry groups (ALL) and calculated for each inferred continental ancestry group separately (AMR, AFR, EUR, SAS, EAS, MID). **B)** Discordance between blood types dervied from O vs non-O tSNPs and those derived from functional variants was plotted along with LD (r^2^) between the O vs non-O functional variant and tSNPs. A Spearman’s correlation coefficient was calculated (-0.97, p-value = 1.51 x 10-38).

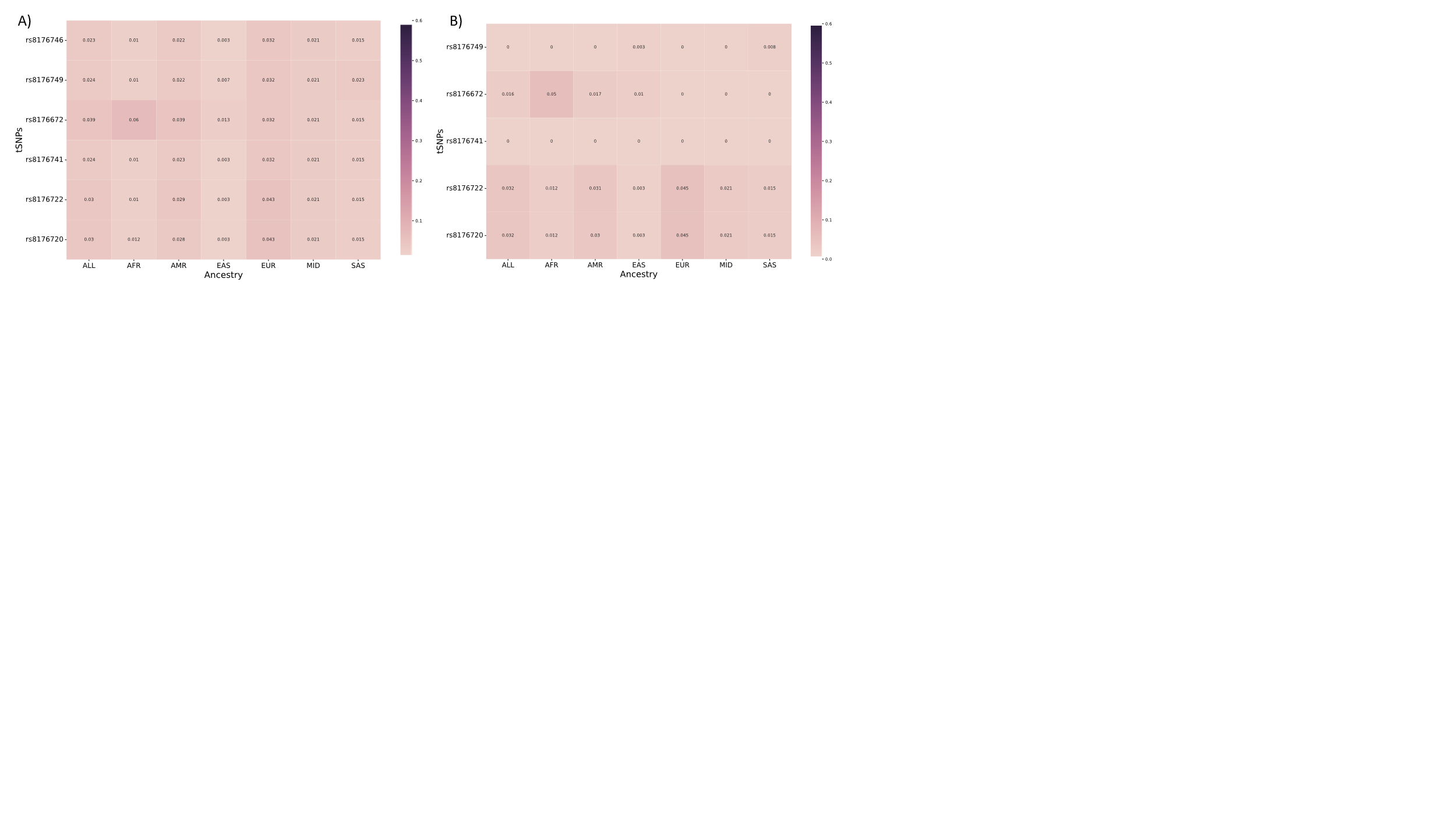

**Supplementary Figure S10.** **Discordance between derivations of ABO blood types**. **A)** ABO blood types were derived with the two functional variants rs8176719, which differentiates between O vs non-O alleles, and rs8176746, which differentiates between A vs B alleles. These were then compared to ABO blood types derived determined by serology. Discordance (proportion of non-matches in a cohort of 10,771) was calculated across all ancestry groups (ALL) and calculated for each inferred continental ancestry group separately (AMR, AFR, EUR, SAS, EAS, MID). **B)** ABO blood types were derived with the two functional variants rs8176719, which differentiates between O vs non-O alleles, and rs8176746, which differentiates between A vs B alleles. These were then compared to ABO blood types derived from the functional variants that differentiates O vs non-O alleles (rs8176719) and tSNPs that differentiate between A vs B alleles. Discordance was calculated across all ancestry groups (ALL) and calculated for each inferred continental ancestry group separately.

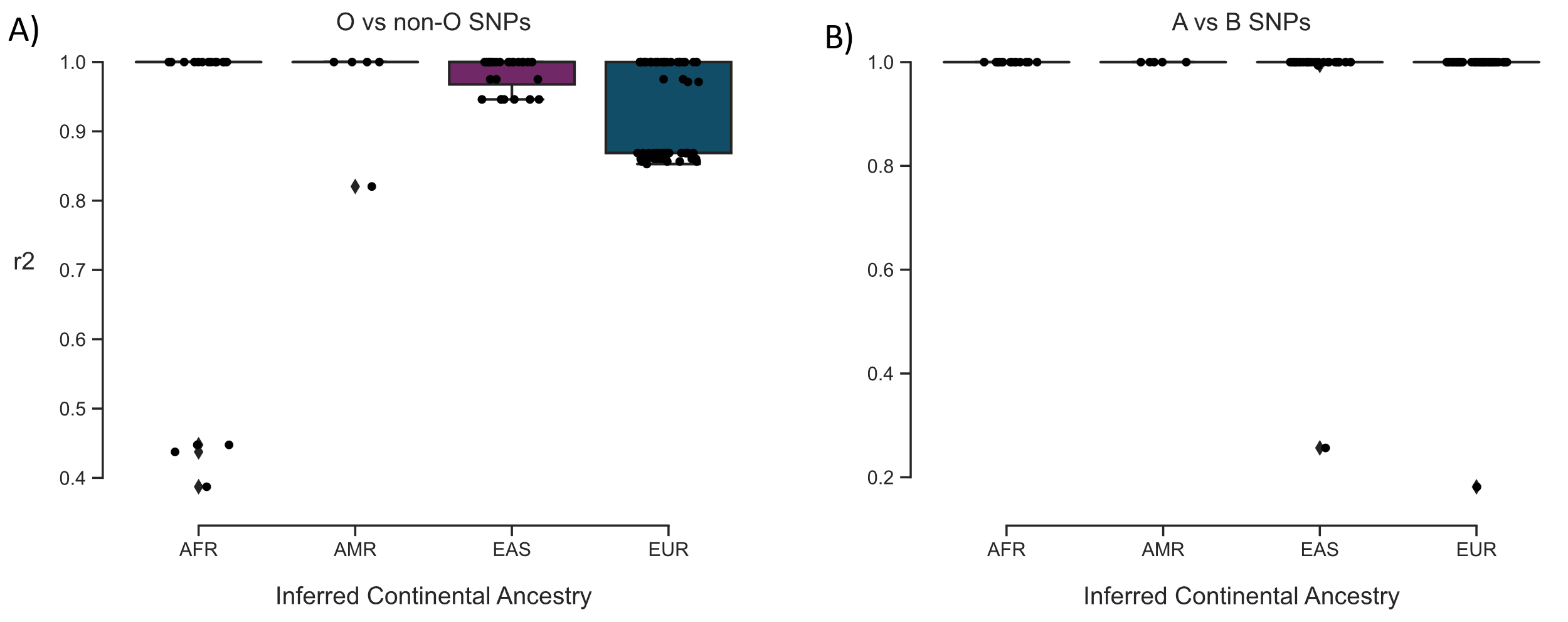

**Supplementary Figure S11.** **Linkage disequilibrium between tSNPs and functional variant for each study.** r^2^ was calculated for each SNP used to determine ABO alleles for each study included in this systematic review. r^2^ was calculated using the reference population (AFR, AMR, EAS, or EUR) that matched the one used by each study. If a study used more than one population, then they are included more than once as each population may have a different r^2^ value. For studies using more than one SNP for a specific population, we assumed they were using the SNP with the highest r^2^ value for their population. The study using an SAS population was not included as they only had an n=1. All SNPs were included regardless of whether they were a tSNP or a functional variant. The r^2^ between a functional variant and itself is 1. We compared the means of the r^2^ values between populations with Dunn’s test of multiple comparisons using rank sums after doing a Kruskal Wallis test.

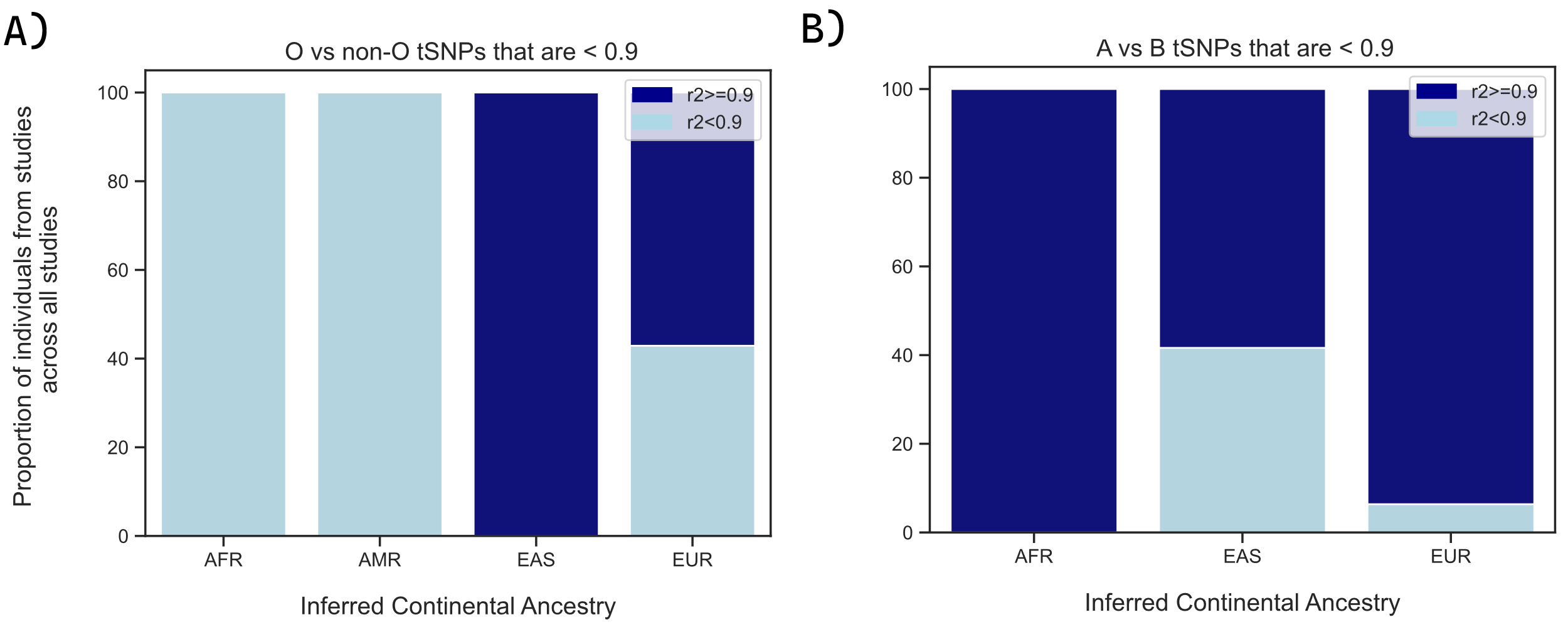

**Supplementary Figure S12. Proportional plot of individuals studies whose ABO alleles were derived with a tSNP that has r^2^<0.9.** We observed the number and proportion of individuals from each population represented in our studies, and evaluated how many were using tSNPs that were poor proxies (r^2^<0.9) for the functional variant. **A)** For a O vs non-O tSNPs, the number of studies with tSNPs with r2 <0.9 are the following: AFR (n=6234), AMR (n=2077), EAS (n=0), and EUR (n=280259). The number of studies with tSNPs r2>0.9 are the following: AFR (n=0), AMR (n=0), EAS (n=133293), and EUR (n=373040). **B)** For a A vs B tSNPs, the number of studies with tSNPs with r2 <0.9 are the following: AFR (n=0), EAS (n=92), and EUR (n=4327). The number of studies with tSNPs r2>0.9 are the following: AFR (n=6151), EAS (n=129), and EUR (n=63594).

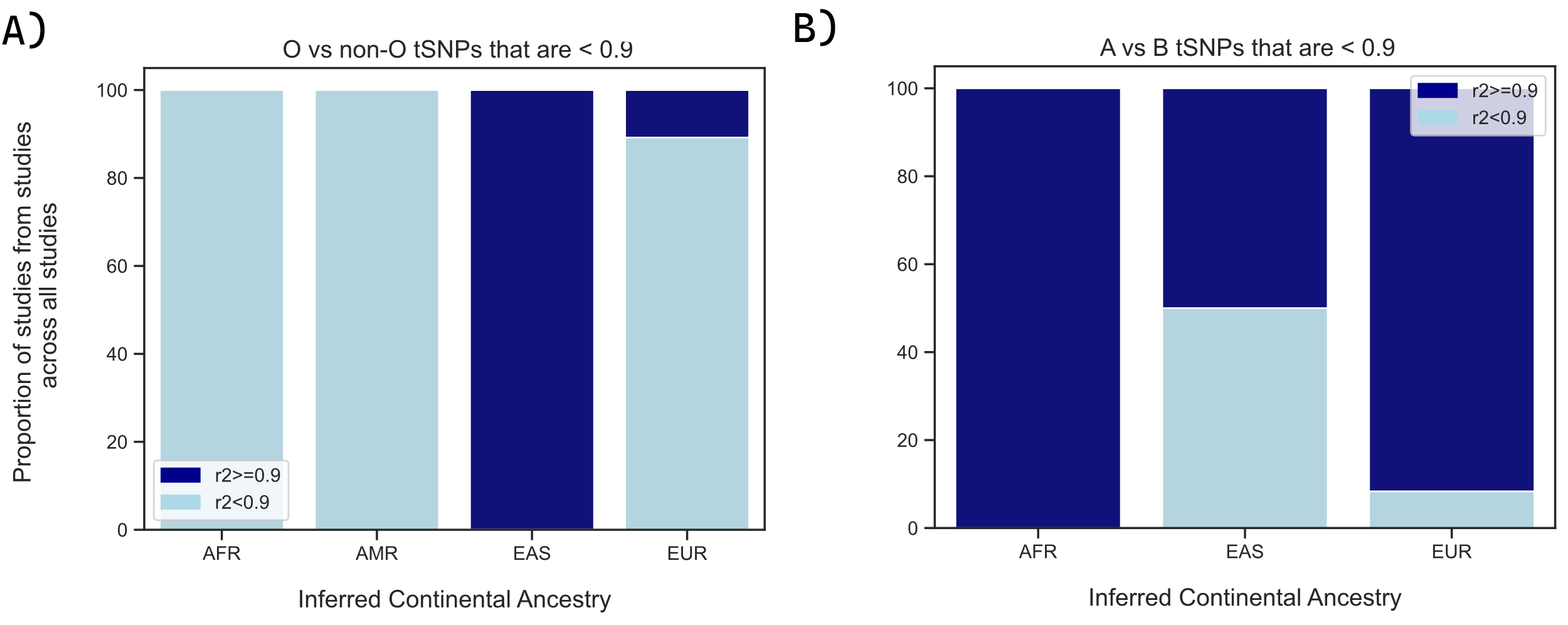

**Supplementary Figure S13.** **Proportional plot of studies that used tSNP with r^2^<0.9**. We evaluated how many studies used tSNPs that were poor proxies (r^2^<0.9) for the functional variant. **A)** For a O vs non-O tSNPs, the number of studies with tSNPs with r2 <0.9 are the following: AFR (n=4), AMR (n=1), EAS (n=0), and EUR (n=33). The number of studies with tSNPs r2>0.9 are the following: AFR (n=0), AMR (n=0), EAS (n=10), and EUR (n=4). **B)** For a A vs B tSNPs, the number of studies with tSNPs with r2 <0.9 are the following: AFR (n=0), EAS (n=1), and EUR (n=1). The number of studies with tSNPs r2>0.9 are the following: AFR (n=3), EAS (n=1), and EUR (n=11)
